# Brain sodium MRI-derived priors support the estimation of epileptogenic zones using personalized model-based methods in Epilepsy

**DOI:** 10.1101/2022.12.14.22283389

**Authors:** Mikhael Azilinon, Huifang E. Wang, Julia Makhalova, Wafaa Zaaraoui, Jean-Philippe Ranjeva, Fabrice Bartolomei, Maxime Guye, Viktor Jirsa

**Affiliations:** Aix Marseille Université, INSERM, Institut de Neurosciences des Systèmes (INS) UMR 1106, Marseille 13005, France; Aix Marseille Univ, CNRS, CRMBM, Marseille, France; APHM, Timone University Hospital, CEMEREM, Marseille, France; APHM, Epileptology and Clinical Neurophysiology Department, Timone Hospital, Marseille, France

**Keywords:** Sodium MRI, Bayesian Inference, Personalized brain network models, Logistic regression, Imbalanced dataset, Drug-resistant focal epilepsy

## Abstract

Epilepsy patients with drug-resistant epilepsy are eligible for surgery aiming to remove the regions involved in the production of seizure activities (the so-called epileptogenic zone network (EZN). Thus the accurate estimation of the EZN is crucial. A data-driven, personalized virtual brain models derived from patient-specific anatomical and functional data are used in Virtual Epileptic Patient (VEP) to estimate the EZN via optimization methods from Bayesian inference. The Bayesian inference approach used in previous VEP integrates priors, based on the features of stereotactic-electroencephalography (SEEG) seizures’ recordings. Here, we propose new priors, based on quantitative ^23^Na-MRI. The ^23^Na-MRI data were acquired at 7T and provided several features characterizing the sodium signal decay. The hypothesis is that the sodium features are biomarkers of neuronal excitability related to the EZN and will add additional information to VEP estimation. In this paper, we first proposed the mapping from ^23^Na-MRI features to predict the EZN via a machine learning approach. Then, exploiting these predictions as priors in the VEP pipeline, we demonstrated that ^23^Na-MRI prior based VEP estimation of the EZN improved the results in terms of balanced accuracy and as good as SEEG priors in terms of the weighted harmonic mean of the precision and recall.

**AUTHOR SUMMARY:** For the first time quantitative ^23^Na-MRI were used as prior information to improve estimation of EZN using the model-based method of VEP pipeline. The priors were based on logistic regression predictions of the EZN, using ^23^Na-MRI features as predictors. The ^23^Na-MRI priors inferred EZNs significantly closer to the clinical hypotheses - in terms of balanced accuracy - than the previously used priors or the no-prior condition.

## INTRODUCTION

Epilepsy is a neurological disorder that affects about 1% of the world population, of which approximately 30% are drug-resistant (Picot, Baldy-Moulinier, Daurès, Dujols, & Crespel, 2008). The epileptogenic zone (EZ), corresponding to the cerebral region generating the seizure, might be arduous to locate, and its localization is crucial in refractory epilepsy that requires surgery. Indeed, surgery success is based on the accurate delineation of the EZ, but this area is rarely reduced to a limited brain region (Bartolomei, Wendling, Bellanger, Régis, & Chauvel, 2001), hence the name of “epileptogenic zone network” (EZN) (Bartolomei et al., 2017) used in the following. Great efforts are being made to find objective and quantifiable markers of EZN including interictal markers such as spikes and high-frequency oscillations (HFO), or ictal markers such as the Epileptogenicity index (EI) (Bartolomei, Chauvel, & Wendling, 2008; Scholly et al., 2019). Therefore SEEG recordings are still the gold standard to define EZN, and allows identification of propagation networks (PZ) as well as regions non-involved by electrical abnormalities (NIZ).

A new method using large scale virtual brain models for the estimation of EZN has been proposed (Jirsa et al., 2017; Proix, Bartolomei, Guye, & Jirsa, 2017; H. E. Wang et al., 2022). The virtual epileptic patient (VEP) is a multimodal probabilistic modeling framework, based on Virtual Brain technology. It is based on a personalized whole brain model simulating brain activity and is derived from subject-specific structural connectivity and neural mass models (NMM). The VEP contains modules providing an estimation of the EZN, but also modules virtualizing surgery strategies (H. E. Wang et al., 2022). The EZN estimation via VEP has been validated not only while modeling seizures recorded with stereotactic-electroencephalography (SEEG) (Jirsa et al., 2017; Makhalova et al., 2022; H. E. Wang et al., 2022) but also on synthetic data (Hashemi et al., 2020; Vattikonda et al., 2021). In addition, the VEP has been compared in a retrospective study of 53 patients to EI type quantification methods and to clinical analysis showing encouraging performances (Makhalova et al., 2022). The VEP provides the first fully nonlinear system analysis of whole brain NMM and works on the whole brain source spaces rather than the sensor recording spaces alone. Here the NMM is Epileptor and was developed by Jirsa, Stacey, Quilichini, Ivanov, and Bernard (2014). Epileptor is a phenomenological model based on a system of coupled nonlinear differential equations with five state variables. All together, these equations generate epileptic dynamics called seizure-like events (SLEs). The parameter *x*_0_ in Epileptor, the excitability for each brain region, is a key parameter to lead the system switch between the normal and ictal states (Houssaini, Ivanov, Bernard, & Jirsa, 2015). The VEP model inversion has been proposed to estimate the parameters of NMM in order to best fit SEEG recordings data. The intrinsic non-linear dynamics of NMM in addition to a large number of model parameters and observations render this inversion problem challenging. To solve this problem in Bayesian inference framework (Aster, Borchers, & Thurber, 2018; Gelman et al., 2013), the usage of priors is paramount since it ensures efficient exploration of the posterior distribution by constraining the parameter space. Several prior knowledge can be incorporated such as plausible range of model parameters, dynamics of unobserved brain state, MRI lesions or even the clinical hypothesis of EZN for instance. The previous VEP priors were mainly based on SEEG recordings processing, providing the location of the initiation of the seizures (Hashemi et al., 2020; Makhalova et al., 2022; Vattikonda et al., 2021; H. E. Wang et al., 2022). It is important to explore other neuroimaging modalities as an additional knowledge and as the prior to complement and potentially improve the identification of EZN in the VEP. Moreover, to avoid invasive recordings such as SEEG in the future, and to increase the relevance of the VEP, an important objective is to feed it with non-invasive data. Here, we explored the potential contribution of ^23^Na-MRI, complementing a recent study seeking to evaluate the link between ^23^Na-MRI measures and excitability, i.e. *x*_0_.

^23^**Na-MRI** is the only way to non-invasively quantify sodium in the brain in vivo. However, it is a challenge because the sodium signal is weak (Madelin, Lee, Regatte, & Jerschow, 2014). In epilepsy, the first ^23^Na-MRI study performed at 3T in a group of human focal epilepsy showed a significant increase of total sodium concentration (*TSC*) in EZN compared to propagation zone (PZ) and non-involved zone (NIZ) (Ridley et al., 2017). Nevertheless, *TSC* has limited specificity for epileptogenicity as it likely reflects intracellular and/or extracellular changes as well as differences in cell density or organization.

The quadrupolar interactions of the 3/2 spin of sodium with the electric field gradient of surrounding molecules (Rooney & Springer, 1991) dictate variation in *T*_2_^*^ decay behavior, of which multiparametric investigation has been made with the biexponential fit of the *T*_2_^*^ decay of the sodium MR signal (Ridley et al., 2018). In this paper, we used ^23^Na-MRI at 7T with the enhanced signal-to-noise. The study of quadrupolar interactions gives an indication of the tissue organization and the molecular environment.

Bi-exponential of the *T*_2_^*^ decay enables the characterization of the apparent short fraction sodium concentration (*Na*_*SF*_) and the apparent long fraction (*Na*_*LF*_), which when added together gives the *TSC* (Ridley et al., 2018). In addition, by quantifying the sodium signal fraction with the short *T*_2_^*^ decay component (*f*) this approach may offer a more relevant metric for studying tissue alterations and potentially provide a better link between sodium homeostasis and neuronal excitability in human epilepsy. In a recent study an increase of *f* in the EZN compared to controls and to PZ and NIZ has been reported, whereas TSC was increased in all regions including PZ and NIZ (Azilinon et al., 2022).

We hypothesized that ^23^Na-MRI data can provide complementary knowledge to the VEP through prior. Thus in this paper, we systematically evaluate the added value of ^23^Na-MRI information to improve the performance of VEP for estimation of EZN. In order to study the individual patterns and to find common features between the different patterns, we combined the different sodium features via machine learning approaches using classification models to classify and predict if a region is epileptogenic or not. In the present study, we exploited the resulting predictions as priors in the VEP framework.

## RESULTS

### *VEP workflow with* ^23^*Na-MRI prior*

The VEP workflow, in Figure 1, starts from clinical imaging (anatomical and diffusion MRI) and electrophysiology (SEEG) data to estimate the EZN via a whole brain neuronal modeling. Briefly, the brain network model is formed by nodes defined by the regions of the VEP atlas linked by the structural connectivity, obtained from the patient-specific imaging data. Note that here the network is patient-specific. Epileptor, a phenomenological neural mass model, is then used to simulate seizure-like activity on each brain region. The signals are generated in the source space and then projected onto the sensors, thus obtaining the simulated activity within each channel of the SEEG electrodes, previously localized.

**Figure 1.**
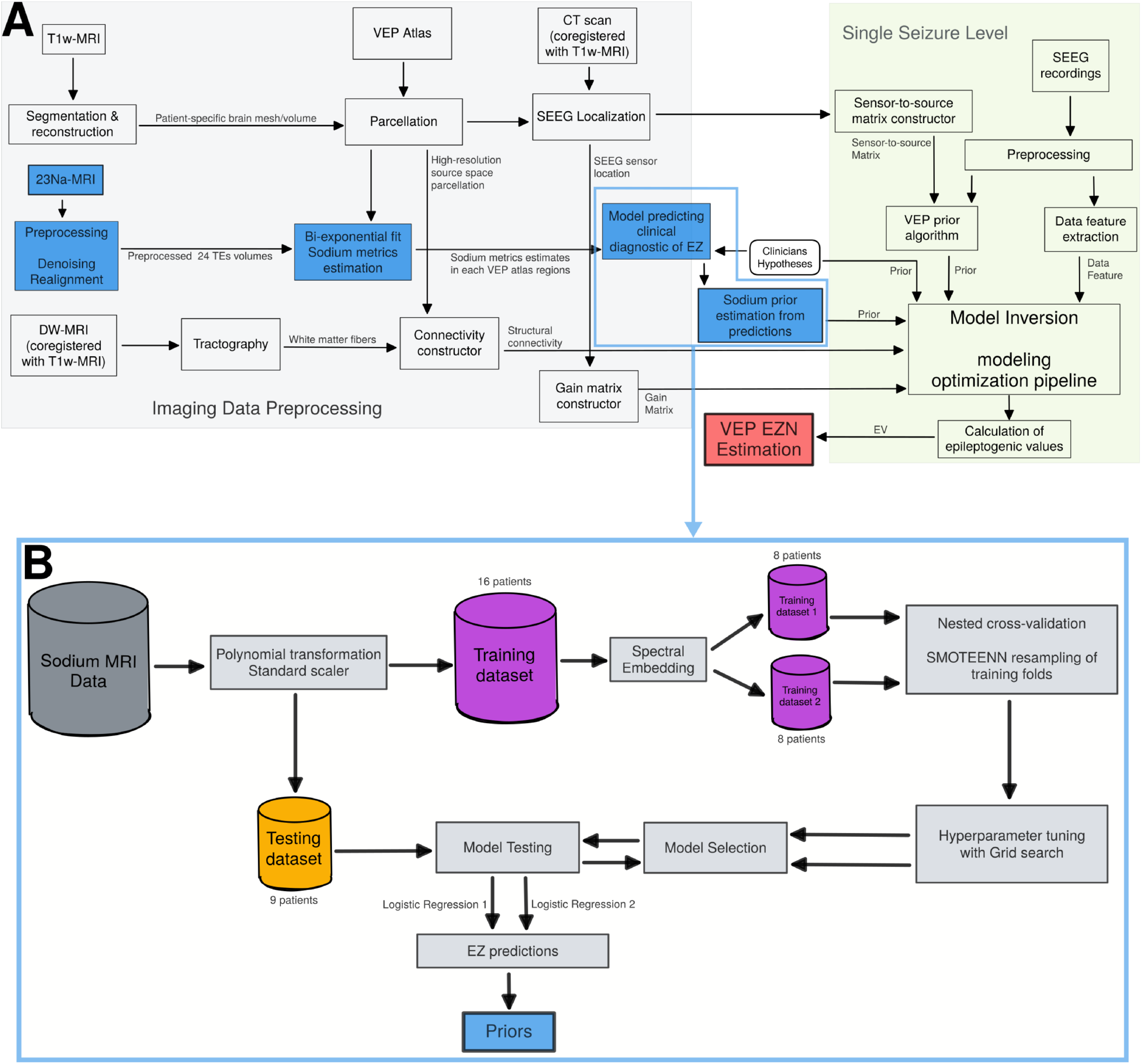
The whole workflow, including (A) flowchart of the VEP pipeline and (B) flowchart of the ^23^Na-MRI based priors’ estimation. A) In the VEP pipeline, the *T*_1_-weighted images defines the patient-specific high-resolution space and are utilized for the parcellation (brain regions on each vertex) according to the VEP atlas. Co-registration of diffusion-weighted images and CT onto *T*_1_-weighted images gives: structural connectivity, gain matrix and sensor-to-source matrix in the patient specific brain space. VEP pipeline is run for each seizure, providing EV distribution as well as the diagnosis, here the goodness of fit for the MAP algorithm. B)^23^Na-MRI volumes at the 24 echo times (TE) are preprocessed before the biexponential fit estimating the ^23^Na-MRI features extracted into VEP atlas ROIs. The quantitative data obtained are transformed and scaled and then train/test split.The training dataset is splitted into 2 subsets according to the spectral embedding first eigenvector values as described in the methods section. Both training dataset were resampled, providing more data points. Hyperparameters were tuned to refine the model selection procedure. Selected models are tested on the testing dataset and the predictions are used as priors.

The model inversion module infers the free parameters of the model from data recording for a personalized simulation. The data features are extracted through SEEG recordings. Here we used the optimization method although a sampling method is also available. The optimization pipeline requires a prior and the likelihood function. Using the L-BFGS algorithm, the goal is to obtain the maximum of the posterior distribution of the model parameters, called **maximum a posteriori (MAP)**. To obtain an EV we bootstrapped 100 MAPs by randomly removing one sensor, which gave us an EV distribution. The regions with the highest EV distribution are labeled as EZN.

Data feature correspond to the power envelope of the signal extracted from the empirical records. We used priors based on ^23^Na-MRI data. For this, we chose to combine the ^23^Na-MRI features via a machine learning model (logistic regression) to predict the EZN. We used the clinical hypotheses as targets in the classification model, to predict these hypotheses with the ^23^Na-MRI features as predictors. The predictions of the two models tested were used as priors (**Na-MRI priors** 1 and 2).

### Feature Importance and Models tuning and selection

The permutation feature importance benefits from being model agnostic and can be calculated many times with different permutations of the feature. Here we estimated the cross-validated permutation importance, with 10 repeats and with balanced accuracy as scoring metric. Balanced accuracy is the accuracy adapted to imbalanced data and is defined as the arithmetic mean of sensitivity (true positive rate) and specificity (true negative rate). While thresholding at 0.05, we obtained 6 features for the training dataset 1 and 8 features for the training dataset 2. The 2 sets of features have 3 common features: *Na*_*LF*_, *TSC* and *f* ^2^. Therefore, we can consider these 3 features as “universal” epileptogenic markers, as they are important independently of the training dataset. The difference in features highlights the fact that each training dataset has a different pattern selective of EZN. Some features, were totally useless, with a poor permutation feature importance score, such as the categorical features “lesional patient” and “lesional zone”, make this information not meaningful to predict EZN with these models.

The selected features are used in the models for hyper-parameters tuning stage. The model selection was based on a cross-validated validation-score higher than 0.65 and a difference score lower than 0.1, resulting in 12 models selected. Next, models were trained on their respective training dataset and provide a (optimized) prediction for each patient. The model with the highest mean testing balanced accuracy, for each training split, was selected for the next stage. The retained parameters for these 2 models are summarized in table 1. Both models get a tolerance C=10, a L2 regularization and similar class weight. On the other hand, solvers differ, logistic regression 1 getting a L-BFGS solver, and logistic regression 2 the SAGA solver.

**Table 1.**
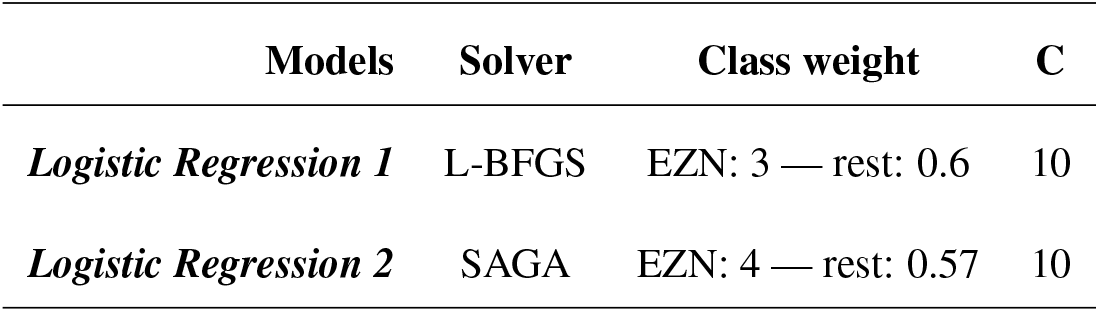
Table of tuned parameters of each selected logistic regression resulting from the grid search procedure.

### Model Testing and Probabilities

Before converting prediction into *x*_0_ and running simulations in the VEP framework, we evaluated performances of the two best logistic regressions, tuned using each training sub-dataset. Models performances were evaluated against clinician’ hypothesis about the EZN. Via graphical analysis (statistical analysis is borderline with few points), we found out that logistic regression predictions are quite robust when using splitted training dataset compared to the three surrogate models with shuffled targets and to models fitted on the whole training dataset without any splitting. This shows that our splitting approach using **spectral embedding** was quite efficient. The performances obtained on models trained with the whole dataset with the correct labels are comparable to those of the surrogate models (Figure 2).

**Figure 2.**
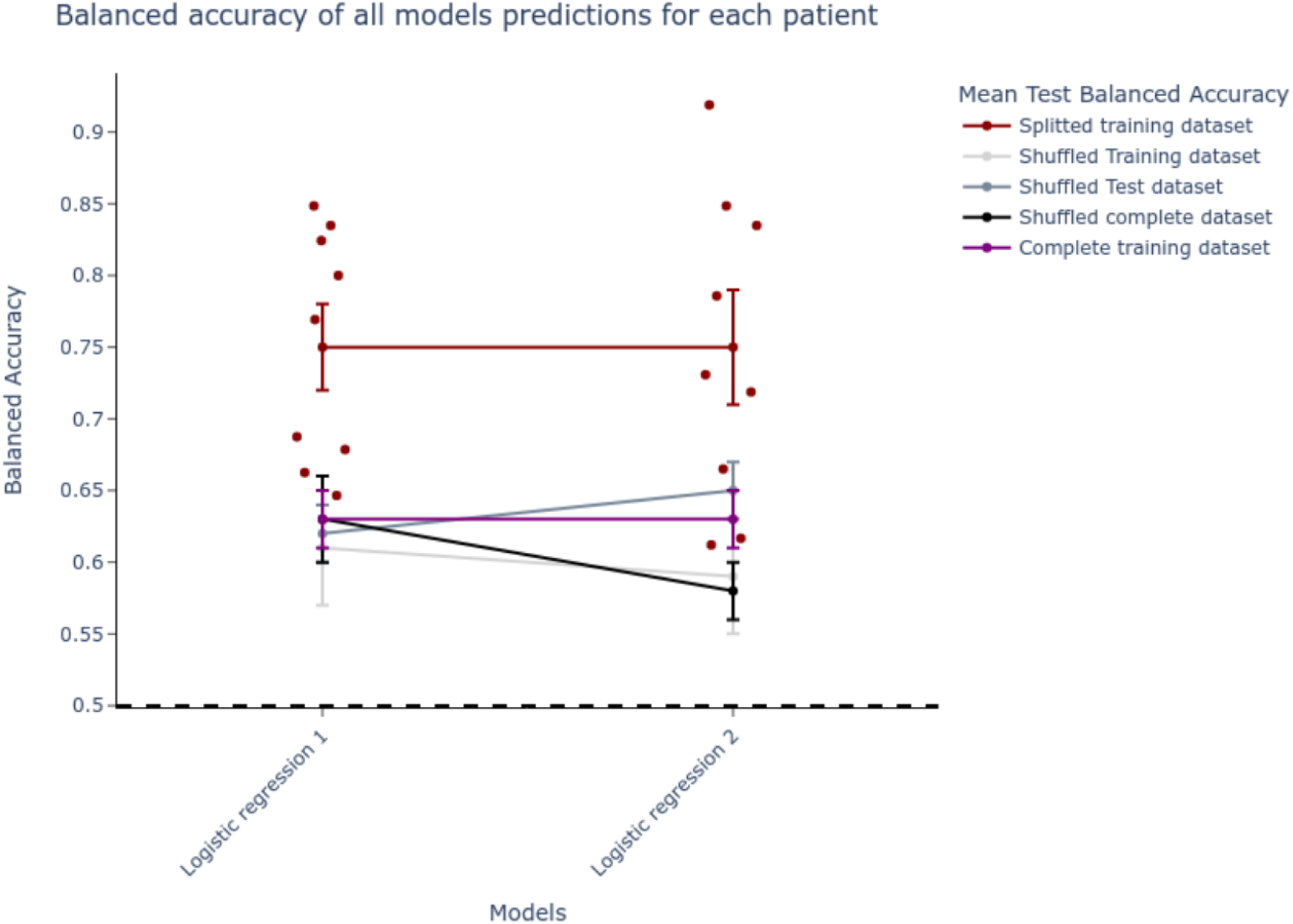
Balanced accuracy of the two selected logistic regression predictions against clinical hypothesis. Logistic regression 1 and logistic regression 2 were tuned using training dataset 1 and training dataset 2 respectively. Red dots represent balanced accuracy of the prediction against clinical hypothesis (i.e. “EZN” or “not EZN”). Dot plus-minus error bars represent the mean balanced accuracy plus-minus standard error of the mean. Splitted training dataset (in red) represents the mean test balanced accuracy for each model trained on the sub-dataset used to tune it. Shuffled means that the target labels of a given sub-dataset were randomly shuffled in the training dataset (light gray), in the testing dataset (dark gray) and in the whole dataset (black). When these models are trained of the complete training dataset, mean balanced accuracy is in purple.

The predictions used to estimate the balanced accuracy were binarized model probability prediction. The threshold was optimized for each model and each patient in order to obtain the best prediction of the EZN. These probabilities are represented in Supplemental Figure 2 for each region’s clinical hypothesis. We observe a clear gradient, with higher probabilities in effective EZN - in the clinical hypothesis - and lower probabilities in NIZ, while PZ probabilities are in between. It is likely that this is due to the predictors, namely ^23^Na-MRI features.

### EZN estimation: Clinical use case

The VEP workflow was applied to empirical data from a female patient in her 20s, with no surgical intervention yet. The patient was initially diagnosed with bilateral temporal plus epilepsy and a radiologically observed bilateral periventricular nodular heterotopia (Supplemental Table 1). The structural connectivity matrix (Supplemental Figure 3) and sensor-to-source mapping (Supplemental Figure 4) were extracted from patient’ *T*_1_-weighted image, diffusion weighted images and post-implantation CT scan data. These matrices, alongside the data features of SEEG seizures recording, were used as input to run the optimization pipeline. Supplemental Figure 5.B. exhibits the distribution of the signal power among all electrodes, alongside one recorded seizure (Supplemental Figure 5.A.). We can observe here a high activity in the left and right anterior hippocampi in the studied seizure. The VEP prior algorithm, by taking into account the onset delay of the seizure in each channel, calculates sensor prior vector based on 52 different frequency bands from 10-110Hz. This prior vector is then projected at source level in two different ways, providing two distinct priors: VEP-M (Supplemental Figure 6), directly maps the prior value of the sensor with the shortest distance to a given source, while VEP-W (Supplemental Figure 7) maps a weighted sum of the prior values of all sensors to each source, based on their distance (see Methods). We also get two other priors from ^23^Na-MRI features. We performed the prediction in the regions investigated by clinicians using SEEG, i.e. predictions were made on the set of VEP atlas regions that were investigated with SEEG and that were included in the EI analysis. Using the ^23^Na-MRI features (*f, Na*_*SF*_, *Na*_*LF*_ and *TSC*) we first trained classification models, logistic regression, in order to predict the diagnosed (clinical hypothesis) EZN (Figure 1.B.). Features were extracted in the considered ROIs, transformed, rescaled and the most important were selected (see Methods). Training dataset is composed of patients that would not be virtualized in the present study, as only patients from the testing dataset would be. Logistic regression was tuned using Grid search function, and after model selection procedure, two logistic regressions (with different hyperparameters and different features) were selected. The predictions made on the testing dataset were used as two distinct priors: ^23^Na-MRI prior 1 and ^23^Na-MRI prior 2. Hence, we ran the optimization pipeline on 6 different prior networks: Na-MRI prior 1, Na-MRI prior 2, VEP-M, VEP-W, uninformative prior (VEP no prior) and clinical hypothesis.

Neurologists (JS and FB) furnished the clinical hypothesis based on the EI index (Bartolomei et al., 2008) and other SEEG data, such as direct electrical stimulations. In this present clinical use case, we showed that ^23^Na-MRI priors complement VEP priors, particularly VEP-W, which together matched the clinical hypothesis. In fact, we were able to retrieve 4 over 6 EZNs (the most evident ones) of the clinical hypothesis, 3 with VEP-W including right and left anterior hippocampi and left posterior hippocampus (Supplemental Figure 8.D.) and 1 (right amygdala) with ^23^Na-MRI prior (Supplemental Figure 8.E and F). This example demonstrates that ^23^Na-MRI is a useful tool to help and complement the diagnosis of the VEP framework.

### Comparison of different priors

At the group level, we analyzed 26 seizures from 9 patients as the test dataset using the VEP pipeline. We used 5 priors for the model inversion on each seizure: Na-MRI-prior 1, Na-MRI-prior 2, VEP-M, VEP-W and VEP-no-prior. Na-MRI-prior 1 and Na-MRI-prior 2 are derived from predictions of the previous section logistic regression 1 and logistic regression 2, respectively. Parameters resulting from model inversion permit simulation proper to each prior. This was also done for the clinical hypothesis of the EZN, using the VEP-EI prior, and the resulting EZN estimation was used as reference in the performance evaluation approach. Evaluation of performance was made with 2 different metrics specifically used while dealing with imbalanced data: balanced accuracy and *F*_0.5_-score (Figure 3). These scores were computed for each prior’ EZN estimation against the EZN estimation of the VEP-EI prior. Bootstrapped paired t-test shows a significantly (*p <* 0.01) higher accuracy of Na-MRI-prior 1 and Na-MRI-prior 2 compared to no-prior. We also can visually see the higher balanced accuracy compared to VEP-M and VEP-W priors, but with lower significance (*p <* 0.05). Bootstrapped paired t-test does not identify any significant difference of *F*_0.5_-score between priors.

**Figure 3.**
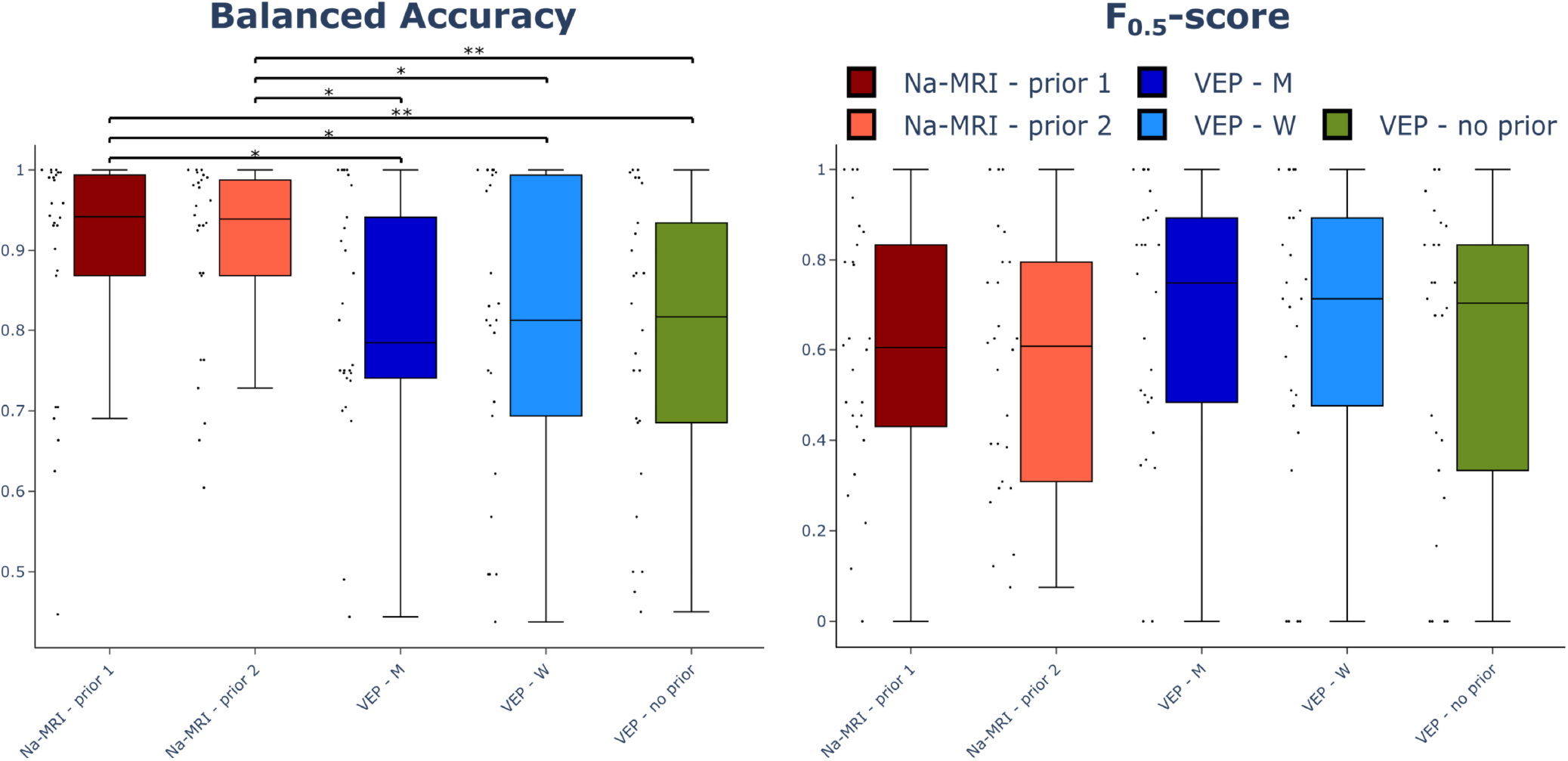
Performance evaluation results. Balanced accuracy (left) and *F*_0.5_-score (right) for each prior EZN estimates for each patient’s seizures against VEP-EI prior EZN estimates. Black dots represent score value for a given seizure on a given patient. Scores of ^23^Na-MRI based priors are represented in shades of red, and those of the currently implemented priors in the VEP (VEP-M and VEP-W) are in shades of blue. Scores of no prior are illustrated with the green box. The top and the bottom of the rectangle in the boxplot represent the first and the third quartile, the line representing the median and the error bars the extrema. * : *p <* 0.05. * * : *p <* 0.01.

## DISCUSSION

For the first time we used quantitative ^23^Na-MRI as additional knowledge to help estimation of EZN using the model-based method of VEP pipeline. The priors were based on logistic regression predictions of the EZN, using ^23^Na-MRI features as predictors. The classification procedure outcomes confirmed the existence of variability in sodium features pattern relative to epileptogenicity, since the use of two training subsamples proved to be more efficient than the use of the whole sample. The procedure provided two models (corresponding to each subsample) built from parameters and features selected specifically for each of them. The predictions of the two models reached an average balanced accuracy of 0.75, much better than those of the surrogate models or the models trained on the whole training set. The ^23^Na-MRI priors inferred EZNs significantly closer to the clinical hypotheses than the currently used priors or the no-prior, in terms of balanced accuracy, but not for *F*_0.5_-score. No significant difference in *F*_0.5_-score reflects no significant difference in precision as *F*_0.5_-score is rather weighted with precision (positive predictive value). On the other hand, balanced accuracy of the prediction, and therefore the sensitivity (true positive rate) and specificity (true negative rate) with each other are significantly improved while using ^23^Na-MRI priors.

### ^23^*Na-MRI features*

The feature engineering methodology makes it impossible to evaluate the contribution of each ^23^Na-MRI feature. Most ^23^Na-MRI studies (the vast majority at 3T) have shown an increase in *TSC* in neurological diseases such as ALS (Grapperon et al., 2019), Huntington’s (Reetz et al., 2012), Parkinson’s (Grimaldi et al., 2021), MS (Maarouf et al., 2017; Zaaraoui et al., 2012) and of course epilepsy (Ridley et al., 2017). Using 7T MRI we also used *f, Na*_*SF*_ and *Na*_*LF*_, measures estimated from the biexponential fit parameters of the 24 TEs (see section ‘Data Processing’). *f* reflects the apparent ratio of short and long *T*_2_^*^ sodium signal decays, and thus encompasses the smallest measurable effects at each TE, with a weighting for short TEs. While in free liquid such as the CSF, the *T*_2_^*^ signal decay is mono-exponential, the quadrupolar interaction of sodium nuclei with the electric field of molecules lead to bi-exponential *T*_2_^*^ decay in the tissues (Berendsen & Edzes, 1973). Here we assumed that *Na*_*SF*_ and *Na*_*LF*_ will be important in the characterization of EZN via logistic regression; in fact both measurements together constitute TSC, hence we have the opportunity to investigate whether or not they can refine the characterization of EZN when not encompassed in TSC, which appear to be crucial for the prediction of the EZN (Supplemental Figure 1). We could therefore imagine in the future, to refine these measures by improving the compartmental models of sodium (multi-exponential decays accounting for intracellular, extracellular, CSF and vascular compartments) for a better characterization of the epileptogenic network.

Together, these measurements provide information on several aspects: (i) sodium homeostasis, (ii) microstructure as the sodium signal may also reflect the structure of the medium in which the sodium is located (Rooney & Springer, 1991), (iii) variation in perfusion (already demonstrated in epilepsy (Kojan et al., 2021; Y. H. Wang et al., 2018)) which may impact blood sodium signal as well, not negligible in the total sodium signal (Driver, Stobbe, Wise, & Beaulieu, 2020). It should also be kept in mind that unlike SEEG, which records electrical activity emanating from neurons, ^23^Na-MRI measures the above-mentioned phenomena in the tissue and thus in neurons and glial cells. Thus, when the SEEG is clearly designated as measuring excitability, we are able to ask ourselves, in view of the results, whether the combination directly measures cortical excitability. We can also ask whether these metrics can be used to analyze phenomena involved in the ongoing epileptogenesis, such as inflammation, glial reaction, plasticity or reorganization (Scharfman, 2007).

### Predictive Model

Being in a supervised training paradigm, we studied the different feature patterns within the EZN in the training dataset. The complete training dataset gave poor performance. Indeed, the raw data showed various patterns, specific to each patient. To provide consistent datasets, we used Spectral Embedding for feature reduction, and we used the eigenvector resultant to split the data into 2 subsamples. In the context of epilepsy, this is easily explained because in our sample, epilepsies are heterogeneous in terms of causes, disease duration and location and anatomical organization. These differences should involve various ionic and metabolic changes in the tissue. Presumably, the patterns of sodium features should vary with these differences. This aspect could not be addressed in this study because of the small number of patients with each type of epilepsy. In the future it may be interesting to explore this issue.

Polynomials and interactions allow the models to learn a more complex decision boundary, thanks to the nonlinearity introduced (Kuhn & Johnson, 2019). However, overfitting becomes a risk and interpreting feature importances gets trickier. In fact, it is difficult to biologically interpret feature combinations because the permutation value of a feature combination is relative to other combinations. All these weaknesses might be addressed in future works. In this work we rather aimed to define an accurate and efficient marker of epileptogenicity than an interpretable model. Nevertheless, in the case of two correlated features, if one of the features is permuted, the model will still have access to the feature through its correlated feature. This will induce a lower importance value for both features, while they may be important. Here, there are indeed correlated features, which expose us to this bias. But we have other downstream strategies to manage the possible biases induced, like the L2 regularization of the models, which will minimize the impact of the ‘useless’ features. In case we missed a feature, we can live with that as long as the model classifies well.

Studying logistic regression predictions probabilities, searching for the optimal threshold for the best performance (balanced accuracy score) we observed that the model seems to be sensitive to clinical hypotheses. In fact there is a gradient of prediction probabilities with EZN the higher, NIZ the lower and PZ in between. This means that although the model was trained on binary targets (EZN vs. non-EZN, i.e. PZ and NIZ), the ^23^Na-MRI features used as predictors show that PZs are not quite NIZs nor quite EZNs. The average probability of EZN is very high (around 0.8), that of NIZ is below 0.4, while that of PZ is slightly higher than NIZ. This is even more pronounced in logistic regression 2 than in logistic regression 1 (Supplemental Figure 2) showing that the model is rather indecisive about PZ. It would be very interesting to study multi-class classification in this context, which would also address the issue of regional variance.

### VEP pipeline and model inversion

The clinical hypothesis used here depict the Epileptogenic zone network (EZN), the propagation zone (PZ) and the non-involved zone (NIZ), but to simplify the procedure we have binarized these assumptions - corresponding to our classification targets - considering a “one vs the rest” strategy, since we are interested in the EZN specifically. This is a strong assumption that affects the choice of *x*_0_ (either -1.5 for a strong excitability or -3 otherwise). Nevertheless, when we look at the probabilities of the models, we notice a gradient, which shows that the models consider the PZs as fuzzy zones between pathological and healthy excitability. It also reflects a continuum of the excitability, referred to in the literature (Bartolomei et al., 2008) We could therefore deepen this study by making a multi-class classification aiming at predicting the EZN, the PZ and the NIZ, to then fix the values of *x*_0_ according to each prediction, with an *x*_0_ between -1.5 and -3 for the propagation zones.

Here we use MAP for the VEP model inversion. The main advantage of MAP is that it is not computationally expensive. However, MAP can get stuck in a local extrema as other optimization algorithms. One of the solutions is to use MCMC-based sampling methods such as NUTS or HMC (Betancourt, 2017; Hoffman, 2014). It was recently shown that HMC sampling implemented in the VEP provides similar results. But in the future, to confirm the results of the present study, or to refine the estimation of EZN, more robust (but also more expensive computationally) techniques like HMC sampling could be employed.

The parameters provided by the optimization procedure using MAP tune the neural mass model, which in turn generate simulated brain activity of a bulk group of neurons. Indeed, reducing 1000 vertices of source activity into one node mapped to a VEP region cancels out the directionality of the current dipole of the folded cortical sheet, which may lead to wrong mapping from sources to sensors and thus possibly introduce errors into the estimation of EZN. Neural Field Model (NFM) might be a solution, simulating brain activity at the vertex level. But tje relatively low resolution of ^23^Na-MRI will complicate the mapping of sodium features onto the vertices. Moreover, the important bias that partial volume effect might introduce in the vertex level sodium estimates will eventually mistake the ^23^Na-MRI based priors. An improvement of the ^23^Na-MRI resolution will facilitate the approach.

Multiple aspects of focal seizures may vary, such as causes, electrographic onset patterns, duration and underlying dynamics. Moreover, it can also happen in a given patient. Our data contains all this possible variability. A priori this variability can be explained by seizure-specific excitabilities (VEP-M and VEP-W) combined with patient-specific structural connectivity. On the other hand, ^23^Na-MRI priors do not take into account the seizure-specific aspect, since only one interictal prior will be used for all seizures of a given patient. Interestingly, this did not have any impact on the outcome, given the high balanced accuracy obtained for ^23^Na-MRI priors. This may mean that commonalities between different seizures could be detected with non-invasive interictal measures such as ^23^Na-MRI, but more studies are needed to make this claim.

### Limitations and Perspectives

In the present study, we based data selection on the clinical hypothesis which lies on SEEG recording analyzes and implantation spatial sampling, considering it as ground truth. As SEEG recordings analyzes may suffer from spatial sampling problems, using it as ground truth is debatable, especially when the ground truth is usually considered to be the brain region which once removed leads to no more seizures (Lüders, Najm, Nair, Widdess-Walsh, & Bingman, 2006). Nevertheless, we had to make a choice for a ground truth using such a prospective database, where the majority of the patients do not have had a surgery, and the choice was to use the best estimation of the EZN common to all patients at the moment of this study. In future, applying a similar approach on seizure-free patients only will be needed to confirm these results.

We have considered a normal distribution of excitability to compute parameter likelihood. The excitability of a region, in the context of a phenomenological model such as Epileptor, is the cumulative sum of the effects of the components that play a role in seizure generation. If these components can be random independent variables then, according to the central limit theorem (Sip et al., 2021), their sum converges to a normal distribution. However, we can imagine that in the case of some epileptogenic lesions, which may or may not generate seizures, a bimodal distribution would be more appropriate.

This last point can also be improved by introducing the regional variance. Currently the parameterization is identical for all the parameters of the model except *x*_0_, and this for all the sources. It would be interesting to vary the parameters according to other biological information, such as cell density, cell type within a region, as well as functional specialization of brain regions. The regional variance can also depend on the functional specialization of regions or the structural connectome.

Regional activity variance has been demonstrated using power spectra and peak frequency of functional data such as SEEG (Frauscher et al., 2018). Most of the anatomical and functional data related to regional variance are available at the group level, making it challenging to use this information in an individualized approach like the one used in this study. In the future it would be interesting to explore how ^23^Na-MRI data can provide information in this sense; now that we are able to extract ^23^Na-MRI data in the whole brain via VEP atlas, we need to determine the right model parameters which can be tuned based on these data to address regional activity variance from the homeostatic point of view.

## CONCLUSION

For the first time, quantitative ^23^Na-MRI was used in the VEP framework. One of the main results of this study is that ^23^Na-MRI features help the VEP to better predict the EZN in terms of a high balanced accuracy when taking the clinical hypotheses as the ground truth. Combining ^23^Na-MRI features via a machine learning approach appears to be a relevant tool to predict the clinical hypothesis and therefore, the derived prediction used as priors in the VEP pipeline can provide more information and a new point of view about the EZN. The next steps would be to upgrade this approach, considering multimodal imaging data, combining other quantitative imaging modalities such proton spectroscopy imaging, as input data to the machine learning pipeline as well as surgery-defined EZN as targets, aiming for more precise estimation of the EZN.

## MATERIALS & METHODS

### Data Acquisition

We obtained the dataset from 25 patients with drug resistant focal epilepsy who underwent a standard pre surgical protocol at La Timone hospital in Marseille. Informed written consent was obtained for all patients in compliance with the ethical requirements of the Declaration of Helsinki and the study protocol was approved by the local Ethics Committee (Comité de Protection des Personnes sud Méditerranée 1).

Patients’ clinical records, neurological examinations, neuropsychological testing, and EEG recordings were assessed in the non-invasive evaluation. The subjects’ clinical data are given in Supplemental Table 1. The evaluation included non-invasive *T*_1_-weighted imaging (see MRI acquisition section of the article for more information) and diffusion-weighted images (DTI-MR sequence, either with an angular gradient set of 64 directions, repetition time = 10.7 s, echo time = 95 ms, voxel size 1.95 × 1.95 × 2.0mm^3^, b-weighting of 1000 s × mm^2^, or with an angular gradient set of 200 directions, repetition time = 3 s, echo time = 88 ms, voxel size 2.0 × 2.0 × 2.0 mm, b-weighting of 1800 s/mm^2^) acquired on a Siemens Magnetom Verio 3T MR-scanner.

In addition, ^23^Na-MRI was acquired using a dual-tuned ^23^Na/ ^1^H QED birdcage coil and a multi-echo density adapted 3D projection reconstruction pulse sequence on a whole-body 7-Tesla Magnetom Step 2 MR system (Siemens, Erlangen, Germany) (for acquisition information see MRI acquisition section of the article). To ensure a sufficient number and distribution of TEs, 3D ^23^Na MRI volumes were obtained at 24 echo times (TEs) ranging from 0.2 ms to 70.78 ms. This approach also takes into account the 5 ms readout of the sequence, needed to acquire ^23^Na signal with short *T*_2_^*^ decay. Signal quantitative calibration into sodium concentration was performed using six tubes (80mm length, 15mm diameter) filled with a mixture of 2% agar gel and sodium at different concentrations: two tubes at 25mM, one at 50mM, two at 75mM and one at 100mM. These external references were positioned in the field of view in front of the subject’s head and maintained using a cap.

Implanted depth electrodes provide patients’ invasive electrophysiological recordings. Electrodes used in SEEG contain 10-18 contacts 2mm long, which are spaced by 1.5mm or 5mm gaps. The SEEG signals were acquired on a 128 channel Deltamed/Natus system with at least a 512 Hz sampling rate and recorded on a hard disk (16 bits/sample) using no digital filter. A high-pass filter (cut-off frequency equal to 0.16Hz at -3dB) was used in the acquisition procedure, as well as an anti-aliasing low-pass filter (cut-off at one third of the respective sampling frequency). SEEG electrodes location are obtained by dint of cranial CT scan, performed after the implantation of electrodes.

### Data preprocessing

To construct the individual brain network models, we performed a volumetric segmentation and cortical surface reconstruction from the patient-specific *T*_1_-MRI data using the recon-all pipeline of the FreeSurfer software package (http://surfer.nmr.mgh.harvard.edu). The cortical surface was parcellated according to the VEP atlas (H. E. Wang et al., 2021) (code available at https://github.com/HuifangWang/VEP_atlas_shared.git).

We used the MRtrix software package to process the DW-MRI (Tournier et al., 2019), using the iterative algorithm described in (Tournier, Calamante, & Connelly, 2012) to estimate the response functions and subsequently used constrained spherical deconvolution (Tournier, Calamante, & Connelly, 2007) to derive the fiber orientation distribution functions. The iFOD2 algorithm (Tournier,, & a. Connelly, 2010) was used to sample 15 million tracts. The streamlines to and from each VEP ROI were assigned and counted, providing the structural connectome. Self-connections, represented by the diagonal of the structural connectome matrix, were excluded by setting them to 0. Finally, the matrix was normalized so that the maximum value was equal to one.

GARDEL (Graphical user interface for Automatic Registration and Depth Electrodes Localization), as part of the EpiTools software package (Villalon et al., 2018), estimated the location of the SEEG contacts from post-implantation CT scans. Afterwards, we coregistered the contact positions from the CT scan space to the *T*_1_-MRI scan space of each patient.

^23^Na-MRI data processing is summarized in Figure 4. All 24 TEs acquired were denoised and realigned on the first TE volume in the same fashion as in the article. VEP atlas volume obtained from VEP pipeline data preparation is ‘voxel cleaned’ via cerebrospinal fluid (CSF) mask obtained with SPM segment function from high resolution *T*_1_ (MP2RAGE) image, after coregistration in the corresponding space. All the voxels shared by this CSF mask and any VEP atlas ROI are erased. This stage was performed in the ultra-high resolution of the 7T *T*_1_ image space (for more information about the MP2RAGE see MRI acquisition section of the article). The cleaned atlas was then projected into ^23^Na-MRI native space for extraction of the 24 TEs signal into each of the 162 VEP atlas ROI. The mean signal of each TEs in each ROI was then fitted with the biexponential model presented in the first part of this manuscript.

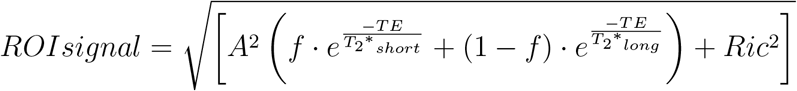

We derived from the biexponential fitting the magnetization (*M* 0) corresponding to the signal fraction estimated by the model in terms of the intercepts of the signal fraction components of the model, obtaining *M* 0_*SF*_ = *A* · *f* and *M* 0_*LF*_ = *A* · (1 − *f*). Then, *Na*_*SF*_ and *Na*_*LF*_ were calculated with raw *M* 0 signal values and the linear fit estimated over the tube phantoms, i.e. slope (a) and intercept (b):

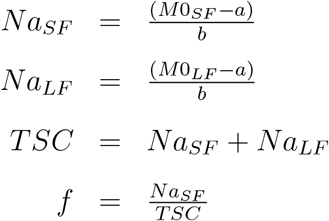

**Figure 4.**
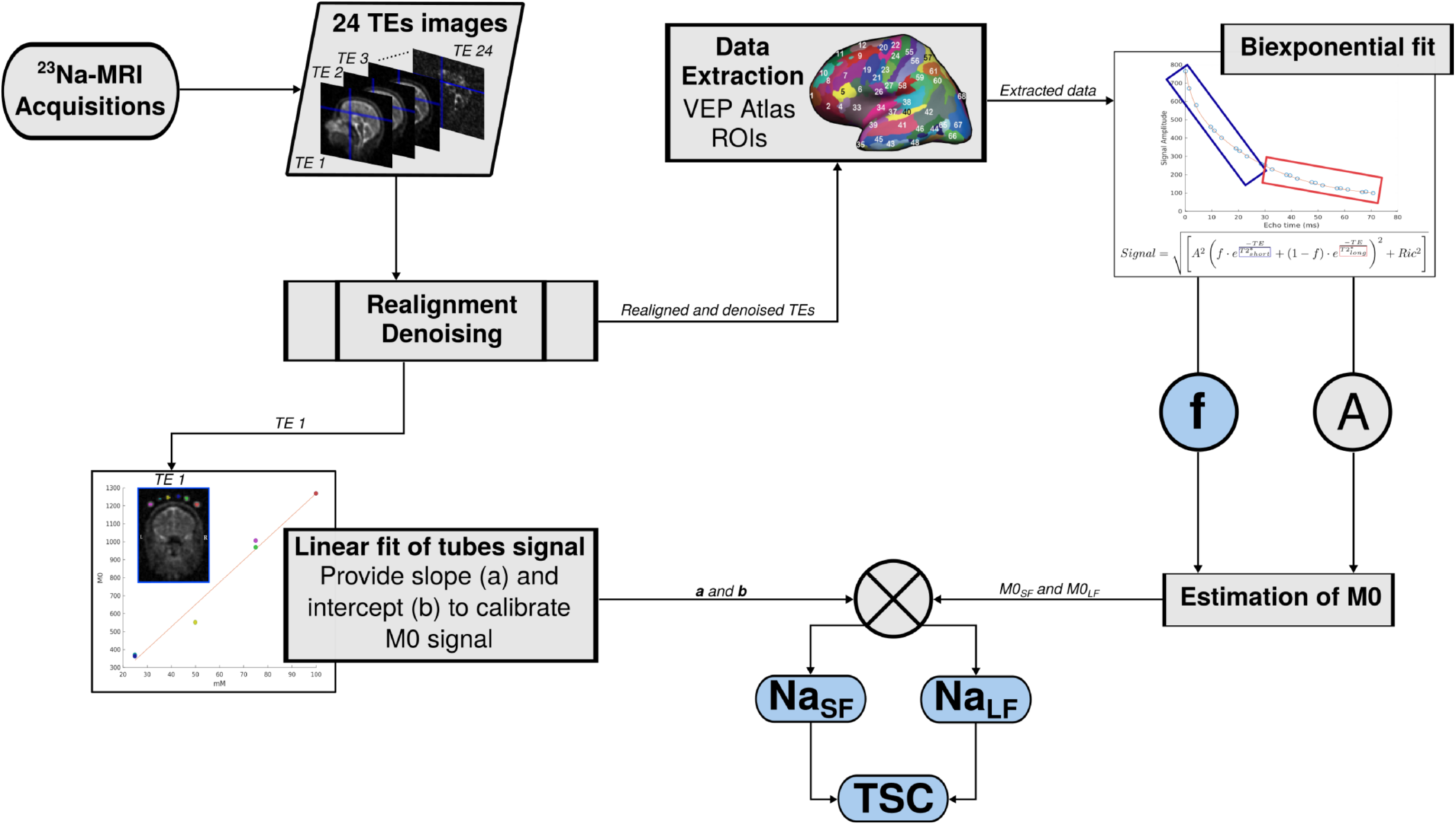
Diagram of ^23^Na-MRI data processing. The 24 volumes acquired with different TEs acquired using a 3D radial density adapted sequence at 7 Tesla are first realigned and denoised. The resulting TE1 image is used for the linear fit of tubes sodium signal to calibrate the sodium signal afterwards. All 24 TEs signal intensities are extracted into VEP atlas ROIs and fitted with the biexponential model. The fit provides parameters such as *A* and *f*, that provide *M* 0 signals fractions. The resulting short and long *M* 0 are calibrated into *Na*_*SF*_ and *Na*_*LF*_. The sum of the resulting *Na*_*SF*_ and *Na*_*LF*_ gives *T SC* (Azilinon et al., 2022; Grimaldi et al., 2021; Ridley et al., 2017).

### VEP model construction

To predict individualized EZNs whole brain neural models (Sanz-Leon, Knock, Spiegler, & Jirsa, 2015) - considering brain areas as nodes connected through edges formed with white matter fibers - was used. Basically, the VEP atlas provides the brain regions, the so called nodes, whereas dWI provides the white matter tracts used to attribute a connection strength to edges. Dynamical equations model the activity of each node, which propagates from one region to another depending on the connection strength between them. The seizure-like activity is modeled with Epileptor, a 6D phenomenological model (Jirsa et al., 2014). The model is composed of 3 neural populations and 3 time scales: fast, intermediate and slow. Taking advantage of time scale separation and using averaging methods, the 6D Epileptor is reduced to a 2D system (Proix, Bartolomei, Chauvel, Bernard, & Jirsa, 2014):

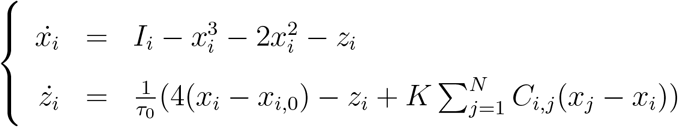

where *τ*_0_ scales the length of the seizure. The state variable *x*_*i*_ describes the activity of neural populations on a fast time scale and can model fast discharges, mostly during ictal periods. The oscillation of the slow permittivity variable *z*_*i*_ drives the system autonomously between ictal and interictal states. The parameter *x*_0_ indicates the degree of excitability (or epileptogenicity) and directly controls the dynamics of the neural population to produce the seizure or not. The coupling between nodes of the network is defined by *C*_*i,j*_, which comes from the structural connectivity. *K* scales the connectivity, which can be varied between simulations to investigate different scenarios. The external input is defined as *I*_1_ = 3.1. In this work we exploited the 2D Epileptor for model inversion in order to speed up the computations essentially.

### Neural mass modeling

Neuronal population macroscopic behavior is modeled by neural masses. In TVB (Sanz-Leon et al., 2015), neural masses represent activity of a single brain region, linked through the structural connectivity, forming a complete brain network model. The global equation for such a model can be given by

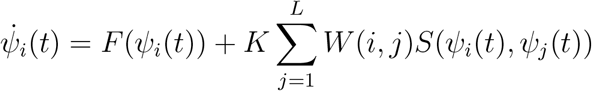

where Ψ_*i*_(*t*) is a state vector of neural activity at brain region *i* and time *t*. 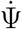 is the temporal derivative of the state vector. *F*, a function of the state, captures the local neural activity; *F* reflects the Epileptor model in the present work. *W* is a matrix of heterogeneous connection strengths between nodes *i* and *j. S* is a coupling function of the local state Ψ_*i*_ and the distant delayed state Ψ_*j*_. The sum across the number of nodes *L* (scaled by a constant *K*) gives the network input received by a node *i*.

### Forward solution

Solving the forward problem and estimating a source-to-sensor matrix permit to map the neural activity from sources (VEP brain regions) to the sensors (SEEG electrods contacts). This matrix *g*_*j,k*_ from source brain region *j* to sensor *k* - also called Gain matrix - is equal to the sum of the inverse of the squared Euclidean distance *d*_*i,k*_ from vertex *i* to sensor *k*, weighted by the area *a*_*i*_ of the vertex on the surface.

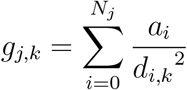

Region *j* contains the vertex *i*, containing a total of *N*_*j*_ vertices. The area *a*_*i*_ of vertex *i* is obtained by summing up one-third of the area of all the neighboring triangles. To obtain the gain for a single region of the brain network model, vertices from the same brain region are summed. The resulting gain matrix has dimensions *M* × *N*, with M being the number of regions and *N* the number of sensors. Matrix product of the simulated activity at the source with the gain matrix gives the simulated SEEG signals.

### Estimation of prior

Here we provide prior to the model inversion module as *x*_0_ parameter values. Prior was usually estimated from SEEG processing (Hashemi et al., 2020; H. E. Wang et al., 2022). The Fourier transform of the SEEG signal provides a spectrogram in 200 consecutive windows. After log transformation of spectrogram values, the seizure onset in each SEEG channel is identified by a sensor-prior vector. The values of the vector are estimated based on channel spectral content across 52 different frequency bands. Each frequency band is defined by the combination of a lower bound ranging from 10 to 90Hz and an upper bound ranging from lower bound + 10 to 120Hz, both in steps of 10Hz. Averaging spectrograms across a given frequency band gives an average time series per SEEG channel, which is thresholded by its 90% percentile. Early increase in a specific frequency band is illustrated by high values of normalized reciprocal value (reciprocal of the first time point above the threshold in each channel). This is considered to be a specific indicator of the SEEG channel where seizure begins. These values are mapped to the source brain region, considering the 60 closest vertices to the sensor for each region. The entries of the sensor-to source-matrix corresponds to

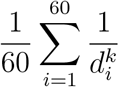

with dik being the euclidian distance from sensor *k* to source vertex *i*. The prior VEP-M consists of assigning the sensor-prior value to the region with the strongest projection to a particular sensor in the sensor-to-source-matrix. VEP-W calculates one onset value per region for each frequency band, which is then averaged across all frequency bands and divided by the max. For both strategies, regions with the resulting value higher than 0.5 are assigned with a *x*_0_ = − 1.5 (putting a high epileptogenicity on nodes representing those regions), and a *x*_0_ = − 3 when lower than 0.5, considering these brain regions as non epileptogenic. In fact, all brain regions get this *x*_0_ value for VEP-no-prior. For the VEP-EI prior, regions diagnosed as EZN by clinicians are assigned with a *x*_0_ = − 1.5 and the other regions with *x*_0_ = − 3. Estimation of ^23^Na-MRI derived priors is detailed in the next section.

### ^23^*Na-MRI based EZN prediction as prior*

Seeking to predict the clinical diagnosis of EZN with ^23^Na-MRI features, we evaluated the classification performances of logistic regression with ^23^Na-MRI derived features as predictors. All classification models are binary, where the positive class is “EZN” and the negative class is “not EZN”. Since the data is highly imbalanced (there are much less “EZN” and “not EZN”), a cost-sensitive framework - through the usage of class weights parameter - is necessary to adjust models prediction (Fernández et al., 2018). The “not EZN” label corresponds to the concatenation of PZ and NIZ labels. These labels are mostly diagnosed based on SEEG recordings processing, using the Epileptogenic Index (EI) (Bartolomei et al., 2008). Hence, the classification was focused on the region with one of those labels. While setting the procedure, we observe a huge variability in Na features patterns, making the model performing poorly. We then decided to split the training dataset to deal with this issue. We tuned the models on both training datasets. The resulting tuned models were used to predict the EZN in of the test dataset patients, providing 2 priors for the VEP pipeline. The whole procedure is detailed below. All priors definitions are summarized and illustrated in Figure 5.

**Figure 5.**
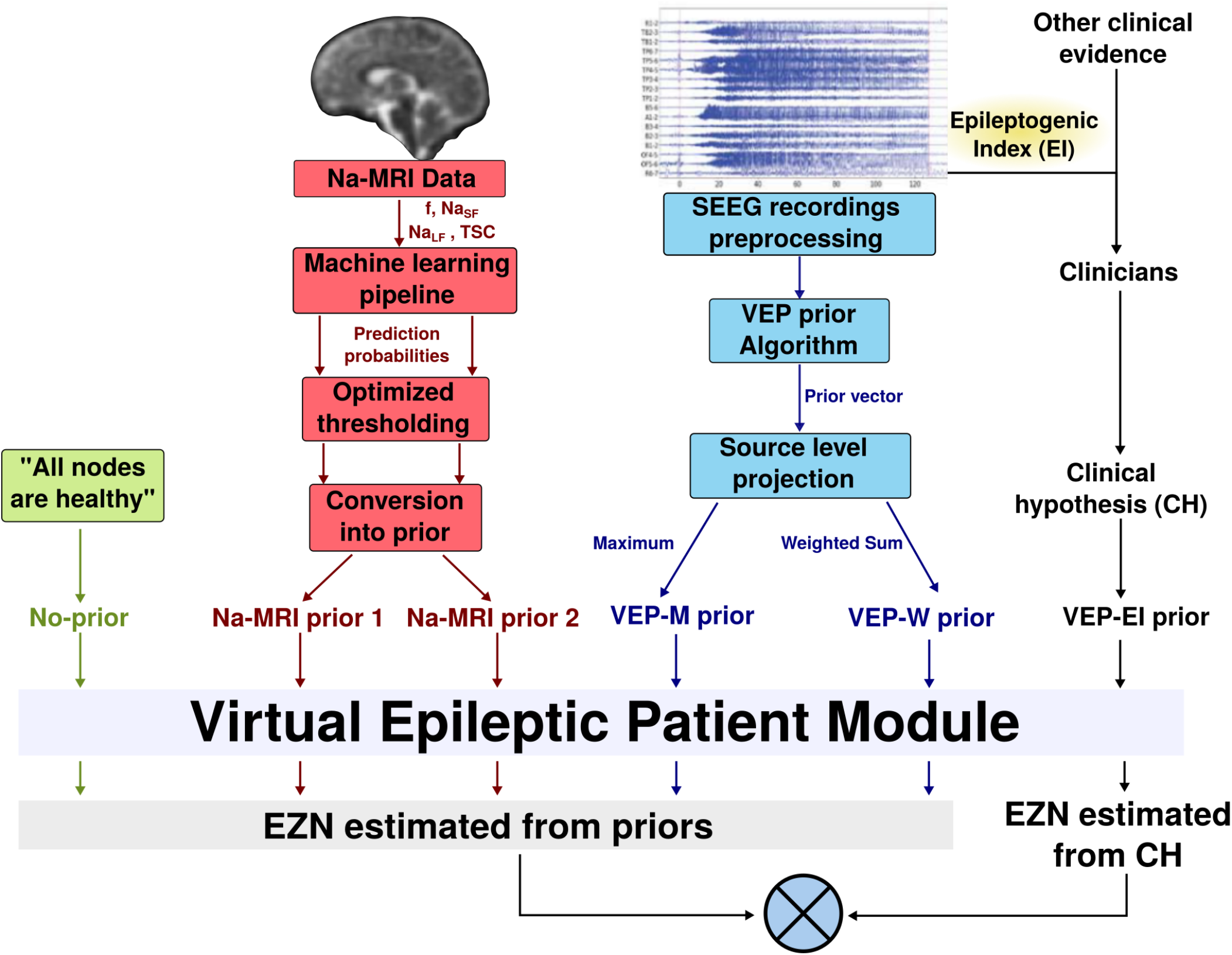
Diagram of the definition of each prior. These procedures are detailed in section *Estimation of prior* and ^23^*Na-MRI based EZN prediction as prior*.

#### Data preparation

Data split The train-test split was performed over the 25 patients dataset, training dataset used only for hyperparameter tuning, model selection and model fitting, and a testing dataset from which we get the predictions of fitted models and use them as priors in the VEP pipeline. Not all of the 25 patients had all the necessary data for the VEP pipeline, so we put those patients into the train dataset. The final training dataset contained 16 patients (9 in the testing dataset).

We observed heterogeneous ^23^Na-MRI feature patterns at the individual level, which initially provided weak performance. So we opted to split the train set to train the models on data with different patterns. We arbitrarily choose to split into 2 different datasets using spectral embedding (Luxburg, 2007) with 2 dimensional projected subspaces. The patients with the mean value in the EZN of the first eigenvector over 0 were included in the training dataset 1, and the others in the training dataset 2, making two sets of 8 patients each. Briefly, spectral embedding is a non linear dimensionality reduction using Laplacian eigenmaps, which preserves the local geometry. Here, we used a graph of nearest neighbors to construct the affinity matrix. After this stage, Laplacian decomposition is applied to the corresponding graph Laplacian. The eigenvectors for each data point provide the resulting transformation.

##### Feature engineering procedure

The first step consists in applying a polynomial transform to features (degree = 2, with interaction terms), basically creating new input features from existing features. The resulting polynomial features correspond to the initial feature plus squared features and interaction terms of each pair of the initial features, obtaining 15 features, in addition to the two categorical features (17 altogether) considered here: “lesional patient”, binary vector containing 1 for regions of a patient with a lesion, and “lesional zone”, binary vector containing 1 for lesional regions. Thanks to the exponent, this transformation separates small and big values and thus changes the probability distribution. The addition of polynomial features to model inputs allowed the model to identify nonlinear patterns alongside linear patterns (Kuhn & Johnson, 2019). Polynomial transformation was followed by a data standardization step, needed for the majority of machine learning models.

The next step was permutation feature importance which corresponds to the decrease in the model score when a feature values are randomly permuted (Breiman, 2001). This procedure can be applied multiple times on repeated permutation of a feature (repeated 10 times here). For each model and on both training sets, this approach was validated using **nested cross validation**. For each cross validation fold, permutation feature importance got a fitted predictive model and training-split of a fold as inputs. Then it computed the reference score of the model on the data, in this instance balanced accuracy. Next, each feature was randomly permuted 10 times, computing at each repetition the permuted score. The importance was finally obtained from the difference between the reference score and the permuted scores.

The nested cross validation and the resampling technique used in the hyperparameters tuning were also used in this procedure; the cross validation providing the training and the validation folds, where the training fold was resampled according to the resampling procedure **SMOTEENN** described in the next section. The so-called permutation importance is estimated by computing the difference between baselines core and permuted score. For each model and training set, the decision threshold of the permutation importance was set to 0.05, considering (arbitrarily) that the most important features should induce a score drop of at least 0.05 when permuted.

#### Model Selection Procedure and priors estimation

Feature permutation outcomes are used as predictors in the following steps. Models hyper-parameters were tuned using a grid search function, which searches over a specified set of parameters values (Table 1) the best mixture of parameters. Each mixture provides a model that will fit on the training dataset, and then provide a performance score, here balanced accuracy. Cross-validated grid search over a parameter grid optimizes parameters of the models. We used a nested cross validation (CV) approach (Wainer & Cawley, 2018), resampling the train-split of each CV fold using SMOTEENN (Batista, Prati, & Monard, 2004). In our nested CV, the outer fold contains 2 patients and the inner fold is composed of 1 patient. For each fold the minority class is oversampled to 60 points while the majority class is oversampled to 180 points before undersampling by ENN, which preserves a relative imbalance, managed by model class weights. The decision about the sets of parameters to use is based on their respective models performance, using the mean cross-validated validation score and the scores difference (mean CV train score - mean CV validation score). The models should have a mean cross-validated score above 0.65 to remove all the models with a performance close to the chance-level. A score difference lower than 0.1 removes all models that overfit, i.e. train score is much higher than the validation score.

The parameters combinations retained provide 12 models. In order to reduce the number of models, we have chosen a model for each training dataset - as all models reached similar performance - considering the mean test balanced accuracy maximum value. The parameters and scores of the selected models are listed in the table.

Model validation consists in comparing model performance on real data compared with surrogate data (shuffled targets, shuffled training dataset targets and shuffled testing dataset targets). This last stage defines the models used for the prediction/priors estimation stage. Predictions are made from prediction probabilities defined by each model. These probabilities are threshold in order to optimize the test score and obtain the best prediction possible from those models. The thresholding binarized the probabilities, in order to be integrated into the VEP framework, and more specifically into the Epileptor model. The predictions are then converted into *x*_0_, the neural mass excitability parameter.

### Optimization MAP pipeline

To infer the epileptogenicity parameter and source time series for each seizure, we apply a Bayesian modeling approach. According to Bayes’ theorem, the posterior probability distribution *p*(*θ* | *y*) of a parameter *θ* given the data *y* is equal to the product of the likelihood *L*(*y* | *θ*) of the data given the parameter and the prior probability distribution *p*(*θ*) of the parameter divided by the marginal likelihood *p*(*y*) of the data.

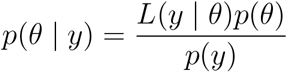

In such a complex multivariate models as (Epileptor) VEP, the marginal likelihood

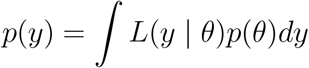

is unsolvable; but as it scales to 1 the integrale across the posterior distribution, one can state the Bayes theorem as

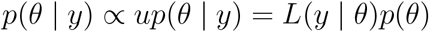

Where the posterior probability is proportional to the unnormalized posterior probability *up*(*θ*|*y*). Normalized and unnormalized posterior distributions have the same properties (namely the shape, maxima and minima) but they are scaled by their respective constant versions. Markov chain Monte Carlo methods generate samples of the unnormalized posterior distribution, hence the posterior distribution can be approximated and inference can be made about the parameters of interest, with a sufficient amount of samples. The maximum-a-posteriori estimate *θ*_*MAP*_ was computed in the optimization pipeline using the L-BFGS (Nocedal, 1980) quasi Newton method.

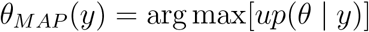

The optimization algorithm performs an iterative process to find *θ*_*MAP*_. It starts with an initial assumption of the parameter before moving through parameter space following the direction of the gradient of the probability distribution. The algorithm terminates after either a maximum of 20,000 steps or convergence has been reached. When changes in parameters, gradients, or probability density between steps are below a certain threshold, convergence is detected. In the current work exploiting the VEP model, the product of prior probability of each parameter and the likelihood of the data provides the posterior probability. Stan (Carpenter et al., 2017) transforms probability into log proba, resulting in a log of posterior proba which corresponds to the sum of log likelihood and log prior probability of all parameters. We specified the prior probabilities for the epileptogenicity parameter *x*_*i*,0_ for brain region *i*, the time scale of the slow variable *τ*_0_, one scaling s and one additive constant of the simulated SEEG, the global coupling scaling factor *K*, and the initial conditions for state variables *x*_*i*_(*t*_0_) and *z*_*i*_(*t*_0_) in region *i*, as well as the distribution width of the extracted data features *ϵ*_*ν*_.

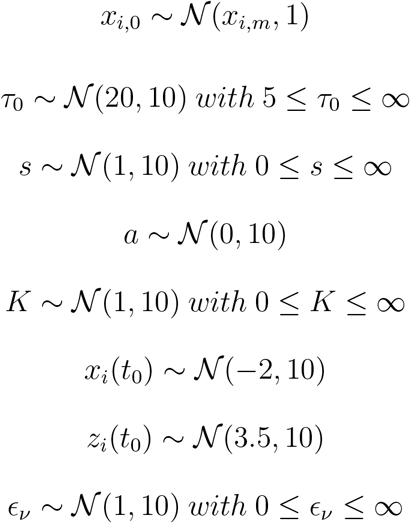

where 𝒩 (*μ, σ*) is a normal distribution with mean *μ* and dispersion *σ* and *x*_*i,m*_ ∈ {− 3, − 1.5} is the epileptogenicity prior for each brain region. Some prior probabilities are truncated by setting a possible minimum value. The likelihood function is given by

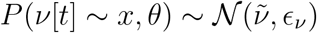

where *ν* and 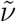 are the empirical and estimated SEEG data features. Both the seizures enveloppes and the simulated SEEG channel power were considered to be the data feature here. Two algorithmic diagnostic metrics are (i) goodness of convergence, the number of runs that terminate properly (the varying of the likelihood converges to the given threshold), and (ii) goodness of fit equals to 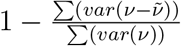, where the sum is across all the SEEG recorded channels.

### Calculation of epileptogenic values

Brain region-specific epileptogenicity values (EVs) are computed based on estimated source time series resulting from the optimization pipeline, just like H. E. Wang et al. (2022). The onset of the seizure *t*_*i*_ in the region *i* corresponds to the first occurence of its source time series (variable *x* of the 2D Epileptor) of region *i* values above a threshold, set at 0 for empirical data. We set *t*_0_ = *min*(*t*_*i*_), *i* = 1, …, 162. When there are no values above 0, meaning no estimated seizure in a brain region, the onset value is set to *t*_*i*_ = 200. We calculate the *EV*_*i*_ of brain region *i*

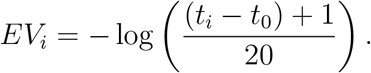

Once the EV vector is normalized to [0, 1] for each optimization run, the optimization pipeline gives the distribution of EVs while considering the sensitivity of the sensor spatial sampling.

### Sensor sensitivity

Measure of estimated EVs confidence was obtained from bootstrapping in the optimization pipeline. The measure evaluates the robustness of the identified source with regard to sensor location sensitivity. The bootstrapping approach relies on a leave-one-out approach, removing one randomly chosen sensor in each bootstrapping sample, whilst the optimization algorithm is run on the remaining data. The procedure is repeated 100 times using a random SEEG contact selection process with replacement, exactly in the same manner that H. E. Wang et al. (2022). All EVs are normalized by subtracting the minimum median EV and dividing by the difference between the maximum and minimum median EVs.

### Statistical Analysis

#### Performance scores

Usually, the performance scores used to evaluate binary classification models, especially when the dataset is imbalanced; they are derived from the confusion matrix. The confusion matrix summarizes the correct and the wrong predictions and thus, helps to understand the number of predictions made by a model for each class, and the classes to which those predictions actually belong. It helps to understand the kind of prediction errors the model made. Figure 3 illustrates this matrix, the derived rates and formula of performance scores used here; true positive rate, false positive rate, true negative rate and false negative rate. Those rates are then used to compute precision (percentage of correct prediction of the positive class) and recall (percentage of correct predictions for the positive class out of all positive predictions).

Balanced accuracy, the imbalanced data adapted accuracy, is the arithmetic mean of sensitivity and specificity (Kelleher, Namee, & D’Arcy, 2015). The classic accuracy score tends to be inflated due to the imbalanced nature of the dataset, which balanced accuracy prevents. The F-beta score used in this project is the harmonic mean of recall and precision Baeza-Yates and Ribeiro-Neto (1999) (Figure 6). As we are more interested in the precision than in recall, we gave more importance to precision than to recall, setting beta = 0.5. These two metrics were first used in combination during parameter tuning procedure as scoring functions, as well as to evaluate VEP pipeline EZN estimation. The model selection is based on balanced accuracy.

**Figure 6.**
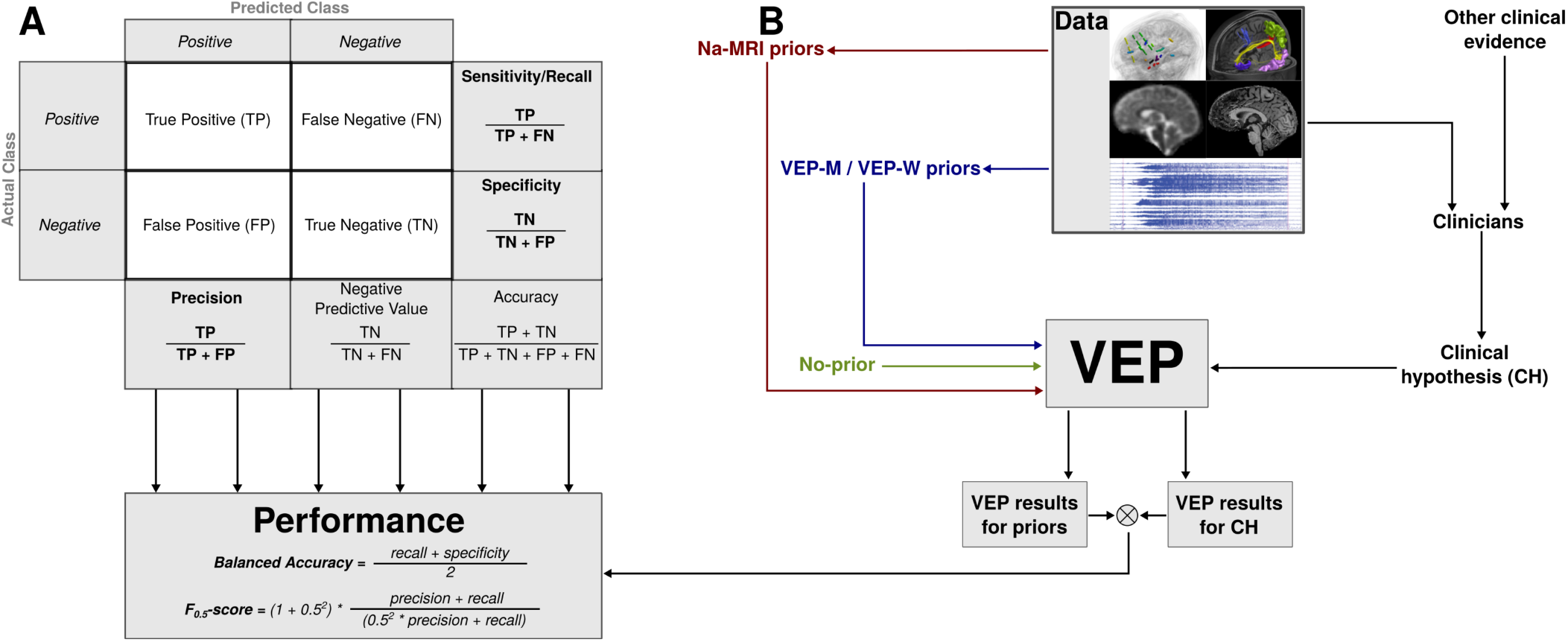
Performance evaluation process. The confusion matrix (A) is used to compute Balanced accuracy and *F*_0.5_-score, the two performance evaluation metrics we explored. Data-driven VEP simulation through the priors (B) provided simulations from the priors and simulation from the CH, confronted altogether to evaluate the performance.

#### VEP outcome analysis

Over the 9 patients of the test dataset that we virtualized using all the described priors (VEP-EI, VEP-M, VEP-W, Na-MRI-prior 1, Na-MRI-prior 2, VEP-no-prior), the VEP pipeline provides results for a total of 26 seizures. In order to evaluate the performance of each prior we compute the balanced accuracy and the *F*_0.5_-score for each seizure, using VEP-EI results as reference. Next, we compared the resulting balanced accuracy and *F*_0.5_-score with a bootstrapped (1000000 resampling) paired t-test comparing each pair of priors. We used the classical threshold of 0.05 for the p-value, also considering the threshold of 0.01.

## Data Availability

The data that support the findings of this study are available on request from the corresponding author. The data are not publicly available due to sensitive information that could compromise the privacy of research participants.

## ACKNOWLEDGMENTS

The authors would like to thank L. Pini, C. Costes, and V. Gimenez for data acquisition and study logistics. We would also like to thank M. Woodman, V. Sip, A. Vattikonda and M. Hashemi for helpful discussions. This work has received support from the French government under the “Programme Investissements d’Avenir”, Excellence Initiative of Aix–Marseille University –A*MIDEX (AMX-19-IET-004), 7TEAMS Chair, EPINOV (Grant ANR-17-RHUS-0004) and ANR (ANR-17-EURE-0029); and from the European Union’s Horizon 2020 Framework Program for Research and Innovation under the Specific Grant Agreement No. 785907 (Human Brain Project SGA2) and No. 945539 (Human Brain Project SGA3).

## SUPPORTING INFORMATION

Supplemental materials can be downloaded here.

## AUTHORSHIP CONTRIBUTION

**Mikhael Azilinon**: Investigation, Conceptualization, Methodology, Data Curation, Formal Analysis, Writing - original draft.

**Huifang E. Wang**: Investigation, Methodology, Data Curation, Formal Analysis, Writing - original draft.

**Julia Makhalova**: Data Curation, Formal Analysis.

**Wafaa Zaaraoui**: Resources.

**Jean-Philippe Ranjeva**: Validation, Writing - review and editing.

**Fabrice Bartolomei**: Funding Acquisition, Resources, Writing - review and editing.

**Maxime Guye**: Funding Acquisition, Conceptualization, Supervision, Writing - review and editing.

**Viktor Jirsa**: Project Administration, Funding Acquisition, Resources, Conceptualization, Supervision, Validation, Writing - original draft.

## TECHNICAL TERMS

^23^**Na-MRI** Imaging technique allowing to quantify human brain sodium in vivo.

**Virtual Epileptic Patient (VEP)** Module simulating epileptic patient’s brain activity to assist clinicians in the identification of the Epileptogenic zones (EZN).

**Epileptor** phenomenological model based on a system of coupled nonlinear differential equations generating epileptic dynamics.

**Neural Mass Models (NMM)** A straightforward approaches to modelling the activity of neuronal populations, the so-called neural masses, described as groups of neurons with a common function and with similar or equal in-going and out-going connections.

**SMOTE** The Synthetic Minority Oversampling TEchnique is an oversampling used for data augmentation of imbalanced dataset.

**SMOTEENN** The combination of over- and under-sampling using SMOTE and Edited Nearest Neighbours (ENN).

**Spectral Embedding** A non linear dimensionality reduction using Laplacian eigenmaps, which preserves the local geometry.

**Nested Cross-validation** Also called double cross validation, this procedure uses two k-fold type loops, meaning that each training fold is also folded.

**Maximum A Posteriori probability (MAP)** Estimate and unknown quantity corresponding to the mode of the posterior distribution.

**VEP priors** Priors providing the location of the initiation of the seizures based on SEEG recordings processing.

**Na-MRI priors** Priors providing the location of the initiation of the seizures based machine learning model (beforehand trained with ^23^Na-MRI) predictions.

## REFERENCES

Aster, R. C., Borchers, B., & Thurber, C. H. (2018). Parameter estimation and inverse problems (2nd editio ed.). Academic Press. doi: 10.1016/C2015-0-02458-3

Azilinon, M., Makhalova, J., Zaaraoui, W., Villalon, S. M., Viout, P., Roussel, T., … Guye, . M. (2022). Combining sodium mri, proton mr spectroscopic imaging, and intracerebral eeg in epilepsy. Human Brain Mapping. Retrieved from https://onlinelibrary-wiley-com.lama.univ-amu.fr/doi/full/10.1002/hbm.26102https://onlinelibrary-wiley-com.lama.univ-amu.fr/doi/abs/10.1002/hbm.26102https://onlinelibrary-wiley-com.lama.univ-amu.fr/doi/10.1002/hbm.26102 doi: 10.1002/HBM.26102

Baeza-Yates, R. A., & Ribeiro-Neto, B. (1999). Modern information retrieval. Addison-Wesley Longman Publishing Co., Inc.

Bartolomei, F., Chauvel, P., & Wendling, F. (2008). Epileptogenicity of brain structures in human temporal lobe epilepsy: A quantified study from intracerebral eeg. Brain, 131, 1818–1830. Retrieved from http://www.ncbi.nlm.nih.gov/pubmed/18556663https://academic.oup.com/brain/article-lookup/doi/10.1093/brain/awn111 doi: 10.1093/brain/awn111

Bartolomei, F., Lagarde, S., Wendling, F., McGonigal, A., Jirsa, V., Guye, M., & Bénar, C. (2017). Defining epileptogenic networks: Contribution of seeg and signal analysis. Epilepsia, 58, 1131–1147. Retrieved from https://onlinelibrary-wiley-com.lama.univ-amu.fr/doi/full/10.1111/epi.13791https://onlinelibrary-wiley-com.lama.univ-amu.fr/doi/abs/10.1111/epi.13791https://onlinelibrary-wiley-com.lama.univ-amu.fr/doi/10.1111/epi.13791 doi: 10.1111/EPI.13791

Bartolomei, F., Wendling, F., Bellanger, J. J., Régis, J., & Chauvel, P. (2001). Neural networks involving the medial temporal structures in temporal lobe epilepsy. Clinical Neurophysiology, 112, 1746–1760. Retrieved from https://linkinghub.elsevier.com/retrieve/pii/S1388245701005910 doi: 10.1016/S1388-2457(01)00591-0

Batista, G. E. A. P. A., Prati, R. C., & Monard, M. C. (2004). A study of the behavior of several methods for balancing machine learning training data. ACM SIGKDD Explorations Newsletter, 6, 20–29. Retrieved from https://doi.org/10.1145/1007730.1007735 doi: 10.1145/1007730.1007735

Berendsen, H. J., & Edzes, H. T. (1973). The observation and general interpretation of sodium magnetic resonance in biological material. Annals of the New York Academy of Sciences, 204, 459–485. Retrieved from https://pubmed.ncbi.nlm.nih.gov/4513164/ doi: 10.1111/J.1749-6632.1973.TB30799.X

Betancourt, M. (2017). A conceptual introduction to hamiltonian monte carlo. Retrieved from https://arxiv.org/abs/1701.02434v2 doi: 10.48550/arxiv.1701.02434

Breiman, L. (2001). Random forests. Machine Learning, 45, 5–32. Retrieved from https://doi.org/10.1023/A:1010933404324 doi: 10.1023/A:1010933404324

Carpenter, B., Gelman, A., Hoffman, M. D., Lee, D., Goodrich, B., Betancourt, M., … Riddell, A. (2017). Stan: A probabilistic programming language. Journal of Statistical Software, 76, 1–32. Retrieved from https://doi.org/10.18637/jss.v076.i01 doi: 10.18637/jss.v076.i01

Driver, I. D., Stobbe, R. W., Wise, R. G., & Beaulieu, C. (2020). Venous contribution to sodium mri in the human brain. Magnetic Resonance in Medicine, 83, 1331–1338. Retrieved from https://onlinelibrary.wiley.com/doi/10.1002/mrm.27996 doi: 10.1002/mrm.27996

Fernández, A., García, S., Galar, M., Prati, R. C., Krawczyk, B., & Herrera, F. (2018). Learning from imbalanced data sets. Learning from Imbalanced Data Sets. doi: 10.1007/978-3-319-98074-4

Frauscher, B., Ellenrieder, N. V., Zelmann, R., Doležalová, I., Minotti, L., Olivier, A., … Gotman, J. (2018). Atlas of the normal intracranial electroencephalogram: Neurophysiological awake activity in different cortical areas. Brain, 141, 1130–1144. Retrieved from https://doi.org/10.1093/brain/awy035 doi: 10.1093/brain/awy035

Gelman, A., Carlin, J. B., Stern, H. S., Dunson, D. B., Vehtari, A., & Rubin, D. B. (2013). Bayesian data analysis, third edition. {CRC} Press. (Google-Books-{ID}: {ZXL}6AQAAQBAJ)

Grapperon, A. M., Ridley, B., Verschueren, A., Maarouf, A., Confort-Gouny, S., Fortanier, E., … Zaaraoui, W. (2019). Quantitative brain sodium mri depicts corticospinal impairment in amyotrophic lateral sclerosis. Radiology, 292, 422–428. doi: 10.1148/radiol.2019182276

Grimaldi, S., Mendili, M. M. E., Zaaraoui, W., Ranjeva, J. P., Azulay, J. P., Eusebio, A., & Guye, M. (2021). Increased sodium concentration in substantia nigra in early parkinson’s disease: A preliminary study with ultra-high field (7t) mri. Frontiers in Neurology, 12, 1610. Retrieved from https://www.frontiersin.org/article/10.3389/fneur.2021.715618 doi: 10.3389/fneur.2021.715618

Hashemi, M., Vattikonda, A. N., Sip, V., Guye, M., Bartolomei, F., Woodman, M. M., & Jirsa, V. K. (2020). The bayesian virtual epileptic patient: A probabilistic framework designed to infer the spatial map of epileptogenicity in a personalized large-scale brain model of epilepsy spread. NeuroImage, 217, 116839. Retrieved from https://www.sciencedirect.com/science/article/pii/S1053811920303268 doi: 10.1016/j.neuroimage.2020.116839

Hoffman. (2014). No u. Journal of Machine Learning Research, 15, 1593–1623. Retrieved from http://mcmc-jags.sourceforge.net

Houssaini, K. E., Ivanov, A. I., Bernard, C., & Jirsa, V. K. (2015). Seizures, refractory status epilepticus, and depolarization block as endogenous brain activities. Physical Review E - Statistical, Nonlinear, and Soft Matter Physics, 91, 10701. Retrieved from https://link.aps.org/doi/10.1103/PhysRevE.91.010701 (Publisher: American Physical Society) doi: 10.1103/PhysRevE.91.010701

Jirsa, V. K., Proix, T., Perdikis, D., Woodman, M. M., Wang, H., Bernard, C., … Chauvel, P. (2017). The virtual epileptic patient: Individualized whole-brain models of epilepsy spread. NeuroImage, 145, 377–388. Retrieved from https://www.sciencedirect.com/science/article/pii/S1053811916300891 doi: 10.1016/j.neuroimage.2016.04.049

Jirsa, V. K., Stacey, W. C., Quilichini, P. P., Ivanov, A. I., & Bernard, C. (2014). On the nature of seizure dynamics. Brain, 137, 2210–2230. Retrieved from https://academic.oup.com/brain/article-lookup/doi/10.1093/brain/awu133 doi: 10.1093/brain/awu133

Kelleher, J. D., Namee, B. M., & D’Arcy, A. (2015). Fundamentals of machine learning for predictive data analytics. {MIT} Press.

Kojan, M., Gajdoš, M., Říha, P., Doležalová, I., Ř ehák, Z., & Rektor, I. (2021). Arterial spin labeling is a useful mri method for presurgical evaluation in mri-negative focal epilepsy. Brain Topography, 34, 504–510. doi: 10.1007/s10548-021-00833-5

Kuhn, M., & Johnson, K. (2019). Feature engineering and selection: A practical approach for predictive models (1st editio ed.). Chapman and Hall/{CRC}. doi: 10.1201/9781315108230

Luxburg, U. V. (2007). A tutorial on spectral clustering. Statistics and Computing, 17, 395–416. Retrieved from https://doi.org/10.1007/s11222-007-9033-z doi: 10.1007/s11222-007-9033-z

Lüders, H. O., Najm, I., Nair, D., Widdess-Walsh, P., & Bingman, W. (2006). The epileptogenic zone: General principles. Epileptic Disorders, 8, S1–9. doi: 10.3109/9780203091708-107

Maarouf, A., Audoin, B., Pariollaud, F., Gherib, S., Rico, A., Soulier, E., … Zaaraoui, W. (2017). Increased total sodium concentration in gray matter better explains cognition than atrophy in ms. Neurology, 88, 289–295. doi: 10.1212/WNL.0000000000003511

Madelin, G., Lee, J. S., Regatte, R. R., & Jerschow, A. (2014). Sodium mri: Methods and applications. Progress in Nuclear Magnetic Resonance Spectroscopy, 79, 14–47. Retrieved from https://linkinghub.elsevier.com/retrieve/pii/S0079656514000211 doi: 10.1016/j.pnmrs.2014.02.001

Makhalova, J., Villalon, S. M., Wang, H., Giusiano, B., Woodman, M., Bénar, C., … Bartolomei, F. (2022). Virtual epileptic patient brain modeling: Relationships with seizure onset and surgical outcome. Epilepsia, 63, 1942–1955. doi: 10.1111/EPI.17310

Nocedal, J. (1980). Updating quasi-newton matrices with limited storage. Mathematics of Computation, 35, 773. Retrieved from https://www.jstor.org/stable/2006193 (Publisher: American Mathematical Society) doi: 10.2307/2006193

Picot, M. C., Baldy-Moulinier, M., Daurès, J. P., Dujols, P., & Crespel, A. (2008). The prevalence of epilepsy and pharmacoresistant epilepsy in adults: A population-based study in a western european country. Epilepsia, 49, 1230–1238. Retrieved from https://onlinelibrary-wiley-com.lama.univ-amu.fr/doi/full/10.1111/j.1528-1167.2008.01579.xhttps://onlinelibrary-wiley-com.lama.univ-amu.fr/doi/abs/10.1111/j.1528-1167.2008.01579.xhttps://onlinelibrary-wiley-com.lama.univ-amu.fr/doi/10.1111/j.1528-1167.2 doi: 10.1111/J.1528-1167.2008.01579.X

Proix, T., Bartolomei, F., Chauvel, P., Bernard, C., & Jirsa, V. K. (2014). Permittivity coupling across brain regions determines seizure recruitment in partial epilepsy. Journal of Neuroscience, 34, 15009–15021. Retrieved from http://www.ncbi.nlm.nih.gov/pubmed/25378166 doi: 10.1523/JNEUROSCI.1570-14.2014

Proix, T., Bartolomei, F., Guye, M., & Jirsa, V. K. (2017). Individual brain structure and modelling predict seizure propagation. Brain, 140, 641–654. Retrieved from https://academic.oup.com/brain/article/140/3/641/2996325 doi: 10.1093/brain/awx004

Reetz, K., Romanzetti, S., Dogan, I., Saß, C., Werner, C. J., Schiefer, J., … Shah, N. J. (2012). Increased brain tissue sodium concentration in huntington’s disease - a sodium imaging study at 4t. NeuroImage, 63, 517–524. Retrieved from https://www.sciencedirect.com/science/article/pii/S1053811912007197 doi: 10.1016/j.neuroimage.2012.07.009

Ridley, B., Marchi, A., Wirsich, J., Soulier, E., Confort-Gouny, S., Schad, L., … Zaaraoui, W. (2017). Brain sodium mri in human epilepsy: Disturbances of ionic homeostasis reflect the organization of pathological regions. NeuroImage, 157, 173–183. Retrieved from https://www.sciencedirect.com/science/article/pii/S1053811917304809 doi: 10.1016/j.neuroimage.2017.06.011

Ridley, B., Nagel, A. M., Bydder, M., Maarouf, A., Stellmann, J. P., Gherib, S., … Zaaraoui, W. (2018). Distribution of brain sodium long and short relaxation times and concentrations: a multi-echo ultra-high field 23na mri study. Scientific Reports, 8, 4357. Retrieved from http://www.nature.com/articles/s41598-018-22711-0 doi: 10.1038/s41598-018-22711-0

Rooney, W. D., & Springer, C. S. (1991). A comprehensive approach to the analysis and interpretation of the resonances of spins 3/2 from living systems. NMR in Biomedicine, 4, 209–226. Retrieved from http://onlinelibrary.wiley.com/doi/abs/10.1002/nbm.1940040502 (_e_print : https://analyticalsciencejournals.onlinelibrary.wiley.com/doi/pdf/10.1002/nbm.1940040502) doi : 10.1002/nbm.1940040502

Sanz-Leon, P., Knock, S. A., Spiegler, A., & Jirsa, V. K. (2015). Mathematical framework for large-scale brain network modeling in the virtual brain. NeuroImage, 111, 385–430. Retrieved from https://www.sciencedirect.com/science/article/pii/S1053811915000051 doi: 10.1016/j.neuroimage.2015.01.002

Scharfman, H. E. (2007). The neurobiology of epilepsy. Current Neurology and Neuroscience Reports, 7, 348–354. Retrieved from https://www.ncbi.nlm.nih.gov/pmc/articles/PMC2492886/ doi: 10.1007/s11910-007-0053-z

Scholly, J., Pizzo, F., Timofeev, A., Valenti-Hirsch, M. P., Ollivier, I., Proust, F., … Bartolomei, F. (2019). High-frequency oscillations and spikes running down after seeg-guided thermocoagulations in the epileptogenic network of periventricular nodular heterotopia. Epilepsy Research, 150, 27–31. doi: 10.1016/J.EPLEPSYRES.2018.12.006

Sip, V., Hashemi, M., Vattikonda, A. N., Woodman, M. M., Wang, H., Scholly, J., … Jirsa, V. K. (2021). Data-driven method to infer the seizure propagation patterns in an epileptic brain from intracranial electroencephalography. PLoS Computational Biology, 17, e1008689. Retrieved from https://journals.plos.org/ploscompbiol/article?id=10.1371/journal.pcbi.1008689 (Publisher: Public Library of Science) doi: 10.1371/JOURNAL.PCBI.1008689

Tournier, J.-D.,, & a. Connelly, F. C. (2010). Improved probabilistic streamlines tractography by 2 nd order integration over fibre orientation distributions. Ismrm, 88, 2010.

Tournier, J. D., Calamante, F., & Connelly, A. (2007). Robust determination of the fibre orientation distribution in diffusion mri: Non-negativity constrained super-resolved spherical deconvolution. NeuroImage, 35, 1459–1472. Retrieved from https://linkinghub.elsevier.com/retrieve/pii/S1053811907001243 doi: 10.1016/j.neuroimage.2007.02.016

Tournier, J. D., Calamante, F., & Connelly, A. (2012). Mrtrix: Diffusion tractography in crossing fiber regions. International Journal of Imaging Systems and Technology, 22, 53–66. Retrieved from http://onlinelibrary.wiley.com/doi/abs/10.1002/ima.22005 (_e_print : https://onlinelibrary.wiley.com/doi/pdf/10.1002/ima.22005) doi : 10.1002/ima.22005

Tournier, J. D., Smith, R., Raffelt, D., Tabbara, R., Dhollander, T., Pietsch, M., … Connelly, A. (2019). Mrtrix3: A fast, flexible and open software framework for medical image processing and visualisation. NeuroImage, 202, 116137. Retrieved from https://www.sciencedirect.com/science/article/pii/S1053811919307281 doi: 10.1016/j.neuroimage.2019.116137

Vattikonda, A. N., Hashemi, M., Sip, V., Woodman, M. M., Bartolomei, F., & Jirsa, V. K. (2021). Identifying spatio-temporal seizure propagation patterns in epilepsy using bayesian inference. Communications Biology, 4, 1–13. Retrieved from https://www.nature.com/articles/s42003-021-02751-5 (Number: 1¡br/¿Publisher: Nature Publishing Group) doi: 10.1038/s42003-021-02751-5

Villalon, S. M., Paz, R., Roehri, N., Lagarde, S., Pizzo, F., Colombet, B., … Bénar, C. G. (2018). Epitools, a software suite for presurgical brain mapping in epilepsy: Intracerebral eeg. Journal of Neuroscience Methods, 303, 7–15. Retrieved from https://linkinghub.elsevier.com/retrieve/pii/S0165027018300955 doi: 10.1016/j.jneumeth.2018.03.018

Wainer, J., & Cawley, G. (2018). Nested cross-validation when selecting classifiers is overzealous for most practical applications. Expert Systems with Applications, 182. Retrieved from https://arxiv.org/abs/1809.09446v1 doi: 10.48550/arxiv.1809.09446

Wang, H. E., Scholly, J., Triebkorn, P., Sip, V., Villalon, S. M., Woodman, M. M., … Jirsa, V. (2021). Vep atlas: An anatomic and functional human brain atlas dedicated to epilepsy patients. Journal of Neuroscience Methods, 348, 108983. Retrieved from https://linkinghub.elsevier.com/retrieve/pii/S0165027020304064 doi: 10.1016/j.jneumeth.2020.108983

Wang, H. E., Woodman, M., Triebkorn, P., Lemarechal, J.-D., Jha, J., Dollomaja, B., … Jirsa, V. (2022). Virtual epileptic patient (vep): Data-driven probabilistic personalized brain modeling in drug-resistant epilepsy. medRxiv, 2022.01.19.22269404. Retrieved from https://www.medrxiv.org/content/10.1101/2022.01.19.22269404v1https://www.medrxiv.org/content/10.1101/2022.01.19.22269404v1.abstract doi: 10.1101/2022.01.19.22269404

Wang, Y. H., An, Y., Fan, X. T., Lu, J., Ren, L. K., Wei, P. H., … Zhao, G. G. (2018). Comparison between simultaneously acquired arterial spin labeling and 18f-fdg pet in mesial temporal lobe epilepsy assisted by a pet/mr system and seeg. NeuroImage: Clinical, 19, 824–830. doi: 10.1016/j.nicl.2018.06.008

Zaaraoui, W., Konstandin, S., Audoin, B., Nagel, A. M., Rico, A., Malikova, I., … Ranjeva, J. P. (2012). Distribution of brain sodium accumulation correlates with disability in multiple sclerosis: A cross-sectional23na mr imaging study. Radiology, 264, 859–867. doi: 10.1148/radiol.12112680

