## Supplemental Data for "Brain sodium MRI-derived priors support the estimation of epileptogenic zones using personalized model-based methods in Epilepsy"

### Supplemental Material

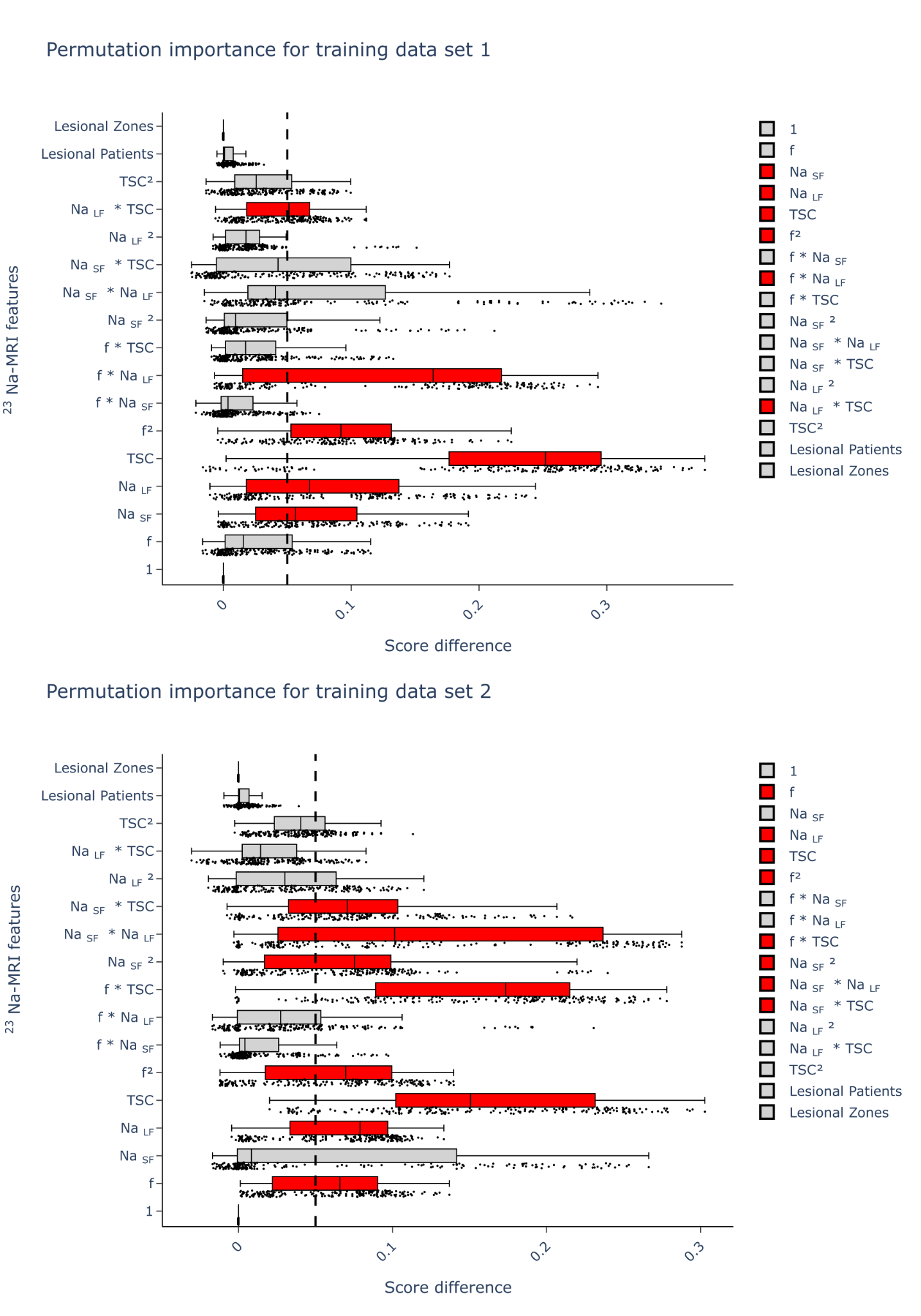

**Figure 1:** Permutation Feature Importance

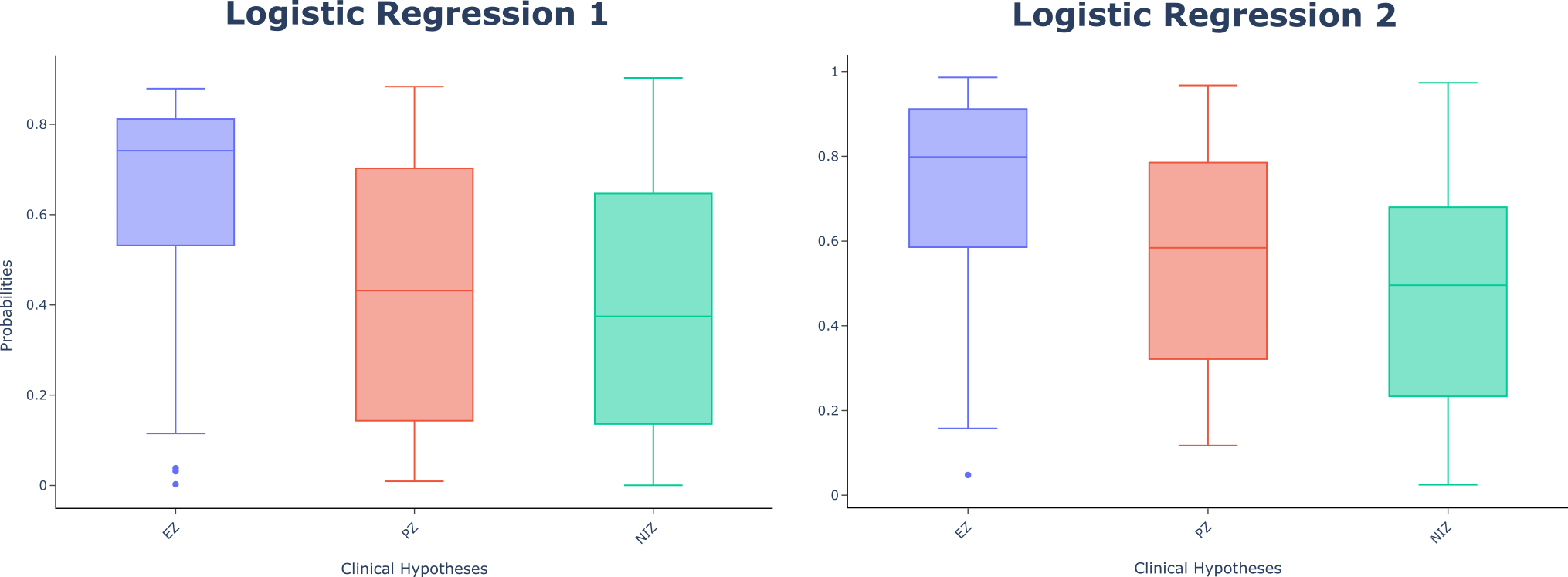

**Figure 2:** Models prediction probabilities per brain region clinical hypothesis

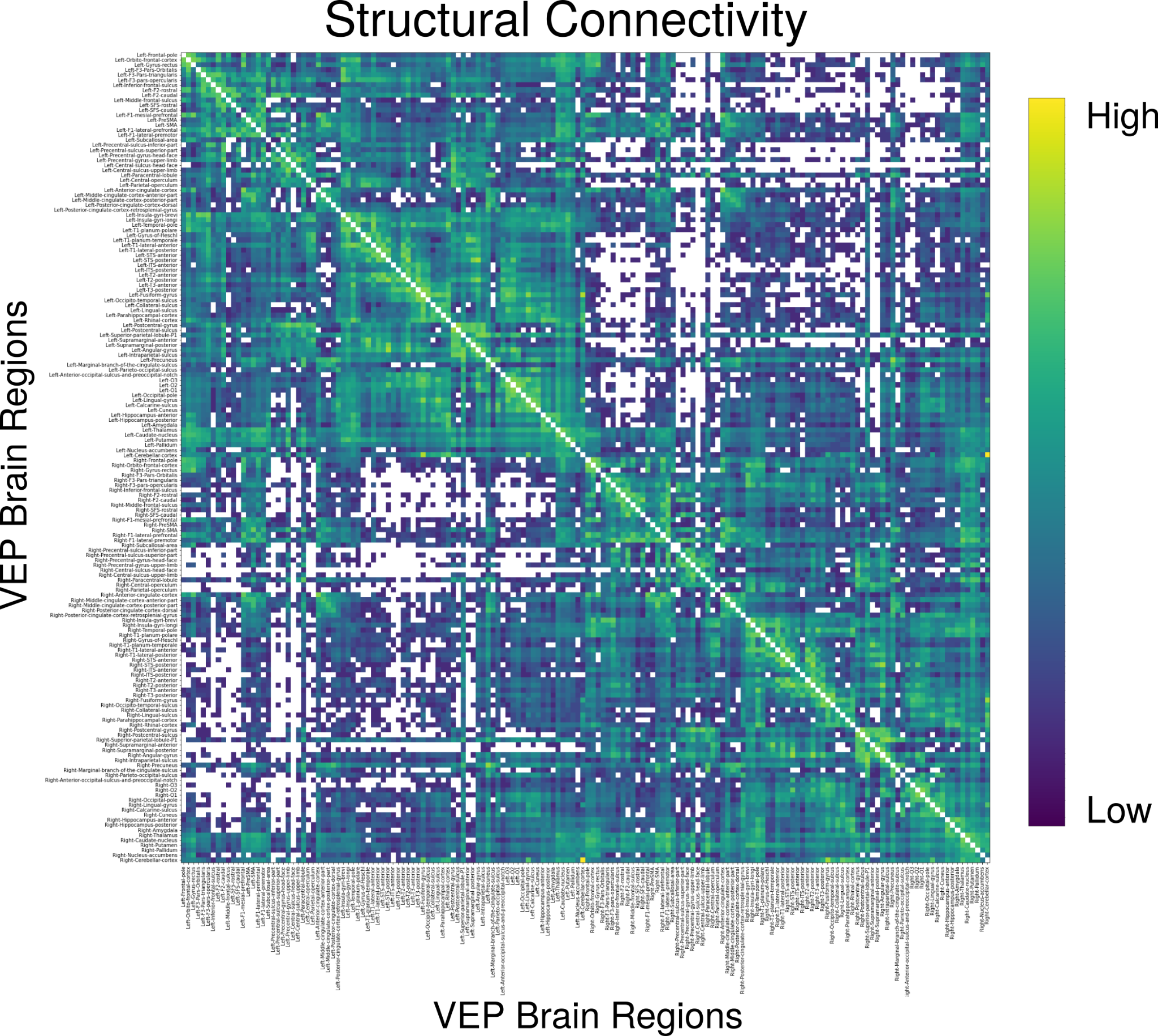

**Figure 3:** Structural connectivity for the clinical case between 162 VEP brain regions.

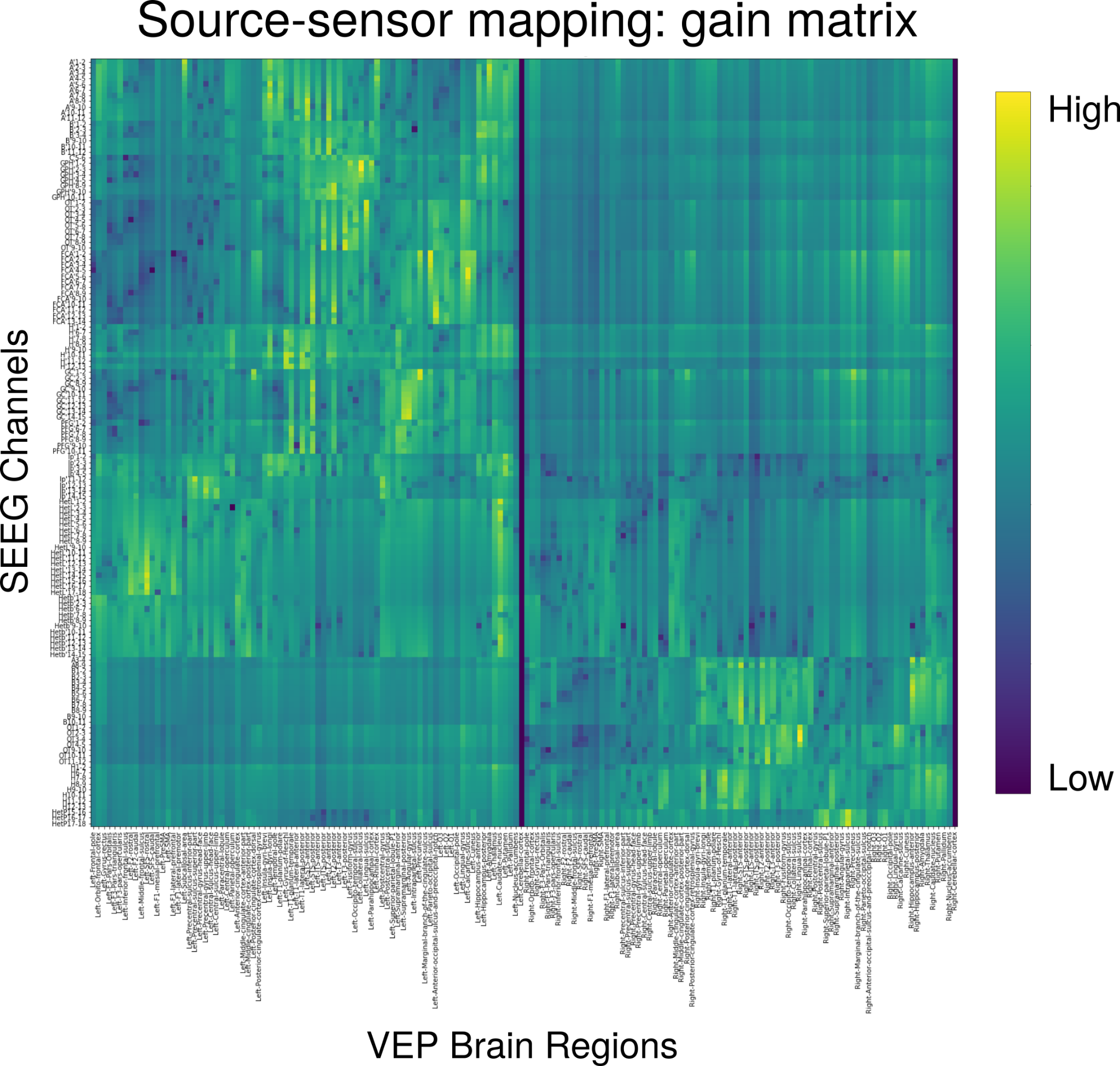

**Figure 4:** Source-to-sensor mapping (gain matrix) for the clinical case between recordings from SEEG bipolar electrodes and 162 VEP brain regions.

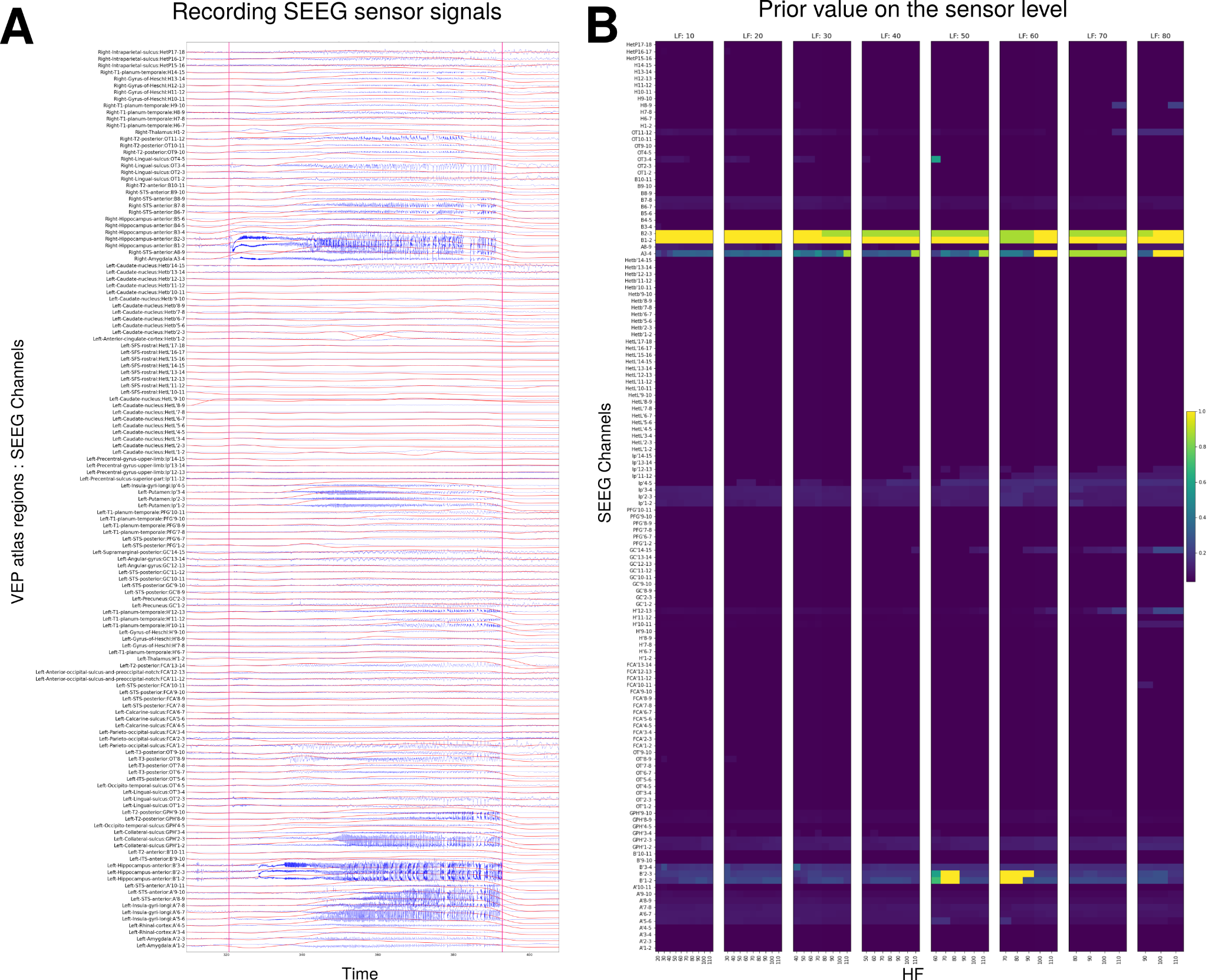

**Figure 5:** (A) Raw time series of the SEEG sensor signals from the example patient (in blue) and the corresponding data feature (in red). (B) The prior values of the sensors on 52 frequency bands; the low frequency boundary is from 10 Hz to 80 Hz with steps of 10 Hz, and the high frequency boundary is from 20 Hz to 110 Hz with steps of 10 Hz.

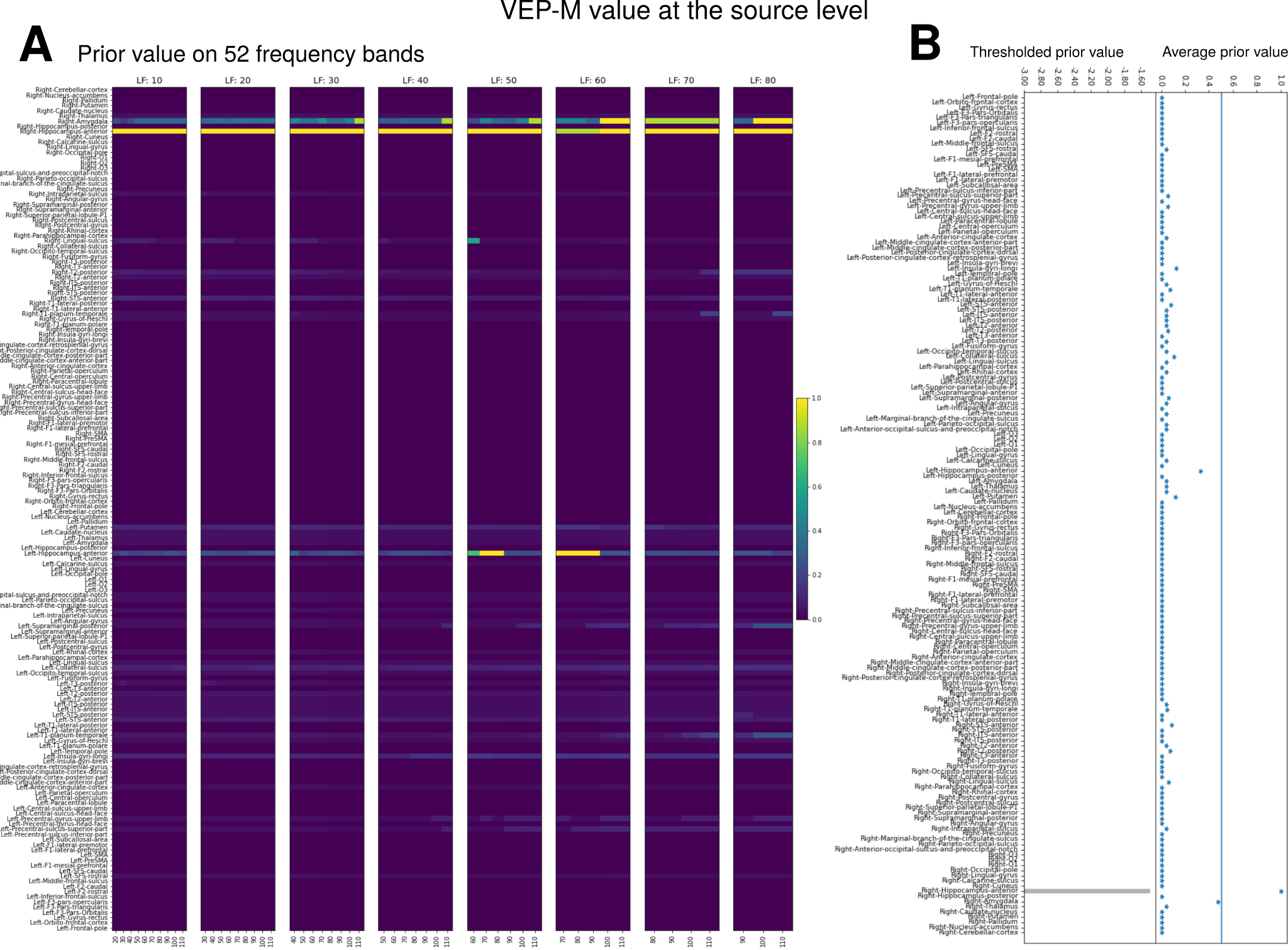

**Figure 6:** Prior value on the source level through the VEP-M algorithm. (A) For each frequency band (as previously defined in Figure 5.B.) sensor prior value is mapped to the closest source (i.e. that has the minimal distance). (B) The bars of the prior values on the left represent the prior values used in the model inversion module for the VEP-M run. The threshold was 0.5. On the right, the stars represent the average across 52 frequency bands of prior values on brain regions.

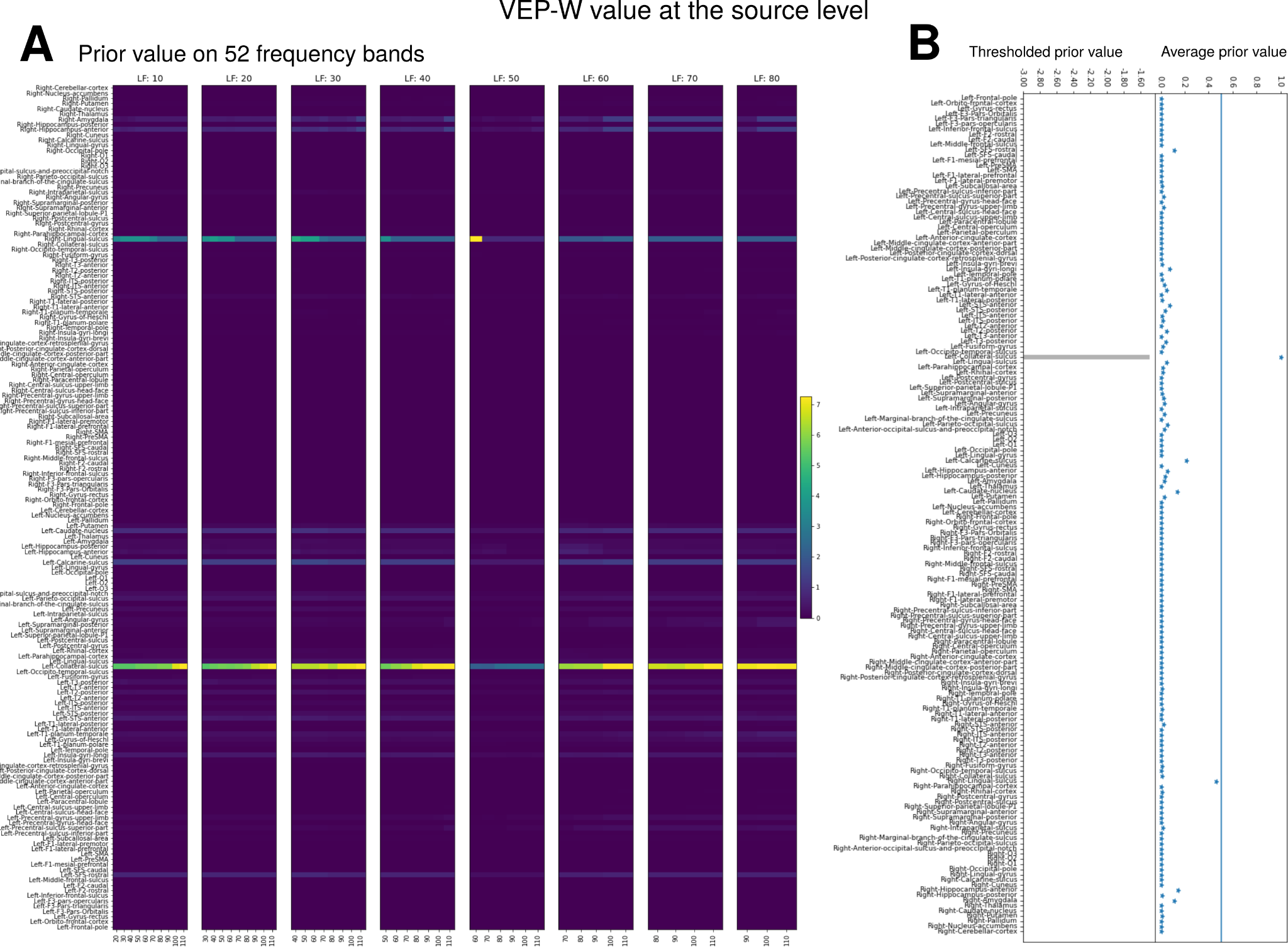

**Figure 7:** Prior value on the source level through the VEP-W algorithm. (A) For each frequency band (as previously defined in Figure 5.B.) weighted sum of all sensors prior value is mapped to each source (i.e. that has the minimal distance). The weight is based on the distance between the source and the sensors. (B) The bars of the prior values on the left represent the prior values used in the model inversion module for the VEP-W run. The threshold was 0.5. On the right, the stars represent the average across 52 frequency bands of prior values on brain regions.

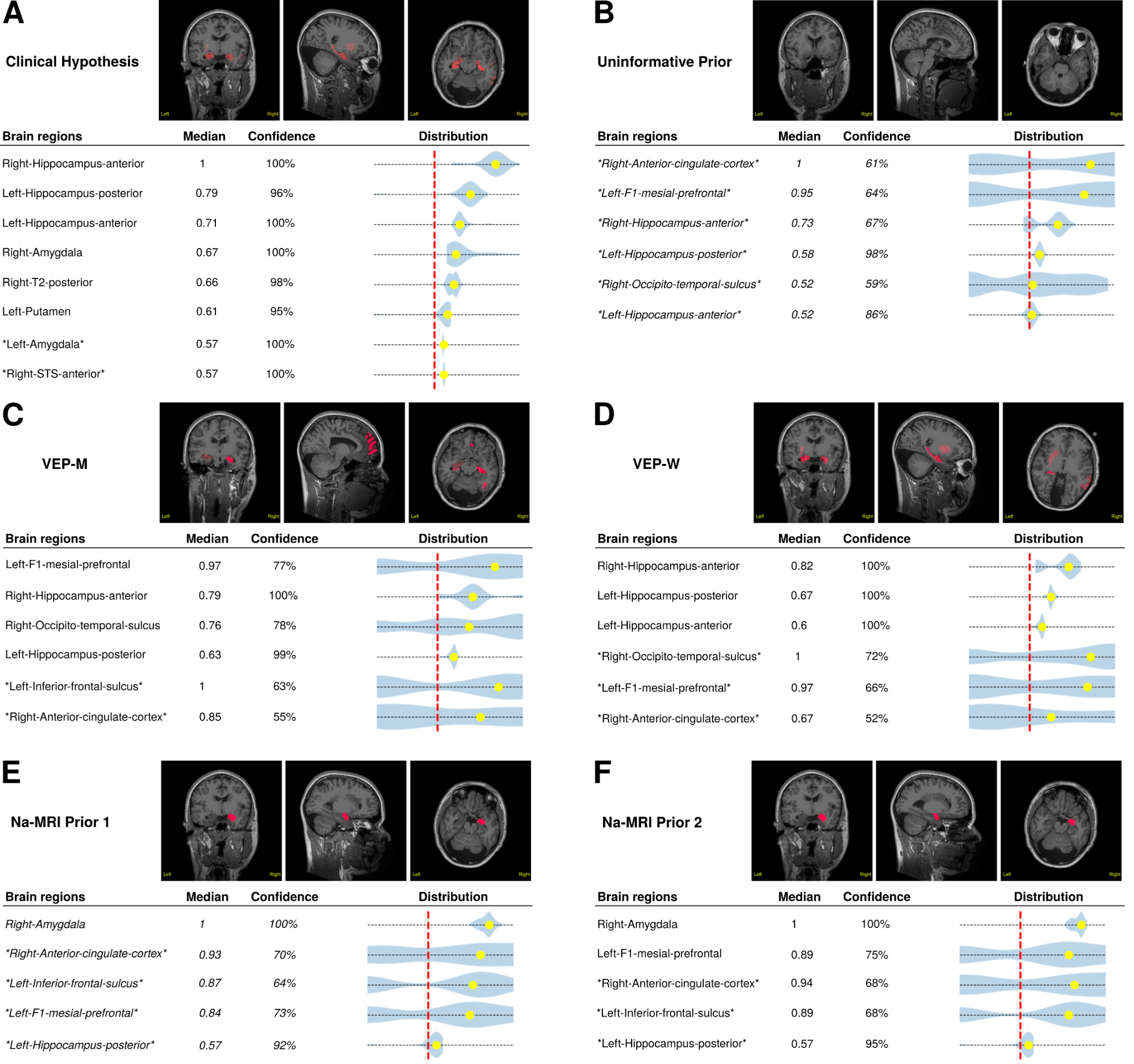

**Figure 8:** The results of 6 VEP runs in an optimization pipeline for the example patient. A, The results of a run using the clinical hypothesis as the prior. B, The results of a run using an uninformative. C, The results of a run using the VEP-M prior. D, The results of a run using the VEP-W prior. E, The results of a run using the Na-MRI prior 1. F, The results of a run using the Na-MRI prior 2. Inside each subfigure, on top, the VEP results are labeled in MRI slides, and on Bottom, the clinical tables, where brain regions are selected when median > 0.6 *and* confidence > 75%.

**Table1: Patients clinical informations**

| **Patient** | **Gender** | **Epilepsy type** | **Side** | **Radiological diagnosis** | **Surgery made** | **Data Splits** |
| --- | --- | --- | --- | --- | --- | --- |
| 1 | F | Temporal latero-mesial | L | Normal | Yes | Training dataset 1 |
| 2 | F | Temporal mesial | R | Normal | Yes | Training dataset 1 |
| 3 | F | Temporal lateral | L | L superior temporal NDT | Yes | Training dataset 1 |
| 4 | M | Temporal mesial | L | Normal | Yes | Training dataset 1 |
| 5 | F | Temporal mesial | R | Normal | Yes | Training dataset 1 |
| 6 | F | Bilateral temporal mesial, R insulo-operculo-frontal | R>L | Normal | No | Training dataset 1 |
| 7 | M | Temporal mesial | R | Normal | Yes | Training dataset 1 |
| 8 | F | Frontal | L | L lateral prefrontal FCD | Yes | Training dataset 1 |
| 9 | F | Temporo-parietal mesial | R | R posterior cingulate FCD | No | Training dataset 2 |
| 10 | M | Insulo-parietal | L | Normal | No | Training dataset 2 |
| 11 | F | Temporo-insular | L | Normal | Yes | Training dataset 2 |
| 12 | M | Temporal mesial | L | Normal | Yes | Training dataset 2 |
| 13 | M | Temporo-insular | L | Normal | No | Training dataset 2 |
| 14 | F | L parietal mesial & bilateral temporal mesial | R&L | L posterior cingulate NDT | Yes | Training dataset 2 |
| 15 | F | Bilateral temporal mesial | R&L | Normal | No | Training dataset 2 |
| 16 | M | Temporal Plus | L | Normal | No | Training dataset 2 |
| 17 | F | Orbitofronto-insular | L | L orbitofrontal FCD | Yes | Testing dataset |
| 18 | M | Temporal latero-mesial | L | Normal | Yes | Testing dataset |
| 19 | M | Insulo-opercular | L | Normal | No | Testing dataset |
| 20 | M | Prefrontal Plus | R>L | Normal | No | Testing dataset |
| 21 | F | Temporal Plus | R | Periventricular nodular heterotopia | Yes | Testing dataset |
| 22 | M | Temporal Plus | R | Normal | Yes | Testing dataset |
| 23 | F | Temporal Plus | L>R | Bilateral periventricular nodular heterotopia | No | Testing dataset |
| 24 | M | Temporal Plus | R | Normal | Yes | Testing dataset |
| 25 | F | Insuli-opercular | R | Periventricular nodular heterotopia | No | Testing dataset |
